# Two-step deep-learning candidemia prediction model using two large time-sequence electronic health datasets

**DOI:** 10.64898/2026.03.03.26347531

**Authors:** Hisato Yoshida, Max W. Adelman, Laila Rasmy, Francis Ifiora, Ziqian Xie, María Alejandra Pérez, Francisco Guerra, Hitoshi Yoshimura, Stephen L. Jones, Cesar A. Arias, Degui Zhi, Masayuki Nigo

**Affiliations:** Center for Infectious Diseases, Houston Methodist Research Institute, Houston, Texas, USA; Department of Dentistry and Oral Surgery, Unit of Sensory and Locomotor Medicine, Division of Medicine, Faculty of Medical Sciences, University of Fukui, Fukui, Japan; Division of Infectious Diseases, Houston Methodist Hospital, Houston, Texas, USA; Weill Cornell Medical College, New York, NY, USA; Division of Pulmonary, Critical Care, and Sleep Medicine, Department of Medicine, Houston Methodist Hospital, Houston, Texas, USA; McWilliams School of Biomedical Informatics, University of Texas Health Science Center at Houston, Houston, TX, USA; Division of Infectious Diseases, Department of Internal Medicine, University of Texas Medical Branch, Galveston, Texas, USA; Center for Health Data Science and Analytics, Houston Methodist Hospital, Houston, TX, USA; Department of Surgery, Weill Cornell Medical College, New York, NY, USA; Department of Infectious Diseases, Faculty of Medical Sciences, University of Fukui, Fukui, Japan

**Keywords:** Candidemia, blood culture, deep learning, empirical antifungal therapy, electronic health record

## Abstract

**Background:** Candidemia is a rare but life-threatening bloodstream infection that remains difficult to predict using conventional risk stratification approaches, highlighting the need for improved predictive strategies. As a result, empiric antifungal therapy is often delayed even in high-risk patients.

**Methods:** We developed a deep learning model (PyTorch_EHR) to predict 7-day candidemia risk by using electronic health record data from two large cohorts (Houston Methodist Hospital System [HMHS] and MIMIC-IV), including adult inpatients who underwent at least one blood culture. Model performance was compared with logistic regression (LR), LightGBM, and established intensive care unit candidemia scores. We further implemented a two-step prediction framework integrating candidemia and 30-day mortality risk models to inform empiric antifungal decision-making.

**Results:** Among 213,404 and 107,507 patients in the HMHS and MIMIC-IV cohorts, candidemia occurred in fewer than 1% (851 [0.4%] and 634 [0.6%], respectively). PyTorch_EHR outperformed LR, LightGBM, and existing candidemia scores, particularly in terms of area under the precision–recall curve (AUPRC) in HMHS and MIMIC-IV. By integrating 30-day mortality risk, the two-step framework identified an additional 20 and 28 candidemia cases beyond the one-step model, increasing coverage to 61% (121/199) and 46% (68/147) in HMHS and MIMIC-IV, respectively. Many patients identified by the two-step framework had high mortality yet did not receive empiric antifungal therapy (61.1% HMHS; 82.6% MIMIC-IV).

**Conclusion:** A two-step deep-learning framework integrating candidemia and mortality risk may support early identification of high-risk patients and facilitate timely empiric antifungal therapy.

Prospective studies are warranted to confirm the findings.

## Introduction

*Candida* spp. account for a minority of bloodstream infections but are associated with high mortality, with overall mortality of approximately 30-40% and attributable mortality of 20–30%.^1–4^ This high mortality is partially explained by the propensity for candidemia to affect patients at high risk of adverse outcomes, including those with critical illness, end-stage renal disease, and malignancy, as well as by delays in empirical antifungal therapy.^5^

Prompt identification of patients with candidemia who may benefit from antifungal therapy remains an important clinical challenge. Blood cultures are the gold standard diagnostic test, but may take several days to return positive for *Candida* spp., as these organisms grow more slowly in blood culture media than bacteria.^6^ Surrogate markers of invasive candidiasis, such as serum 1,3-β-D-glucan, may support diagnosis but are limited by suboptimal specificity and inconsistent availability across institutions, frequently resulting in prolonged turnaround times.^7^ Moreover, 1,3-β-D-glucan–guided antifungal therapy has not been shown to improve clinical outcomes, even when applied to patients traditionally considered at high risk for candidemia.^8^ In the absence of a definitive diagnosis, clinicians must therefore decide whether to initiate empiric antifungal therapy. However, a large randomized clinical trial evaluating empiric antifungal treatment in selected high-risk patients failed to demonstrate a mortality benefit.^9^ Conversely, candidemia may not be considered early in some at-risk patients, and approximately 20–35% of patients with candidemia never receive antifungal therapy^5^, underscoring the difficulty of timely and accurate risk stratification. Several clinical risk prediction models have been developed to aid identification of patients at high risk for candidemia.^10–13^ However, these models have generally focused on narrow subsets of intensive care unit (ICU) populations, limiting their generalizability.

Artificial intelligence (AI)–based machine learning (ML) models offer a potential strategy to support empiric anti-infective decision-making by identifying complex risk patterns within electronic health records (EHRs). ML approaches have previously outperformed traditional logistic regression models for predicting drug-resistant bacterial infections.^14^ Although several studies have applied ML to candidemia prediction^15–18^, their clinical applicability is limited by key methodological concerns. Some models incorporated 1,3-β-D-glucan results as inputs, despite these tests often being unavailable when empiric therapy decisions are made, introducing information leakage.^15^ In addition, some cohorts were artificially balanced to address class imbalance from the rarity of candidemia, and model performance was not evaluated under the true disease prevalence observed in clinical practice.^16–18^ Consequently, the clinical actionability and real-world utility of these models remain uncertain.

We therefore conducted this study to develop deep-learning models that predict candidemia risk among heterogeneous hospitalized patients at the time of blood culture collection—a clinical juncture when empiric antimicrobial and antifungal decisions are often made and AI-based decision support may be most useful. To address the low incidence of candidemia and align with clinical risk stratification, we implemented a two-step framework combining candidemia and mortality prediction models. We compared this approach with existing candidemia prediction scores and with clinicians’ empiric antifungal prescribing patterns across two large electronic health record datasets.

## Methods

### Data Source and Study Population

We performed a retrospective cohort analysis to develop deep-learning models that predict candidemia risk using two large electronic health record (EHR) datasets: the Houston Methodist Hospital System (HMHS) database and the Medical Information Mart for Intensive Care IV (MIMIC-IV, version 3.1). HMHS provides clinical data from eight acute-care hospitals across the Greater Houston area, covering the period from June 2016 to June 2023,^19^ and served as the primary dataset for model training and comparison with traditional machine learning approaches. MIMIC-IV version 3.1, a publicly accessible dataset containing de-identified critical care records from the Beth Israel Deaconess Medical Center in Boston, Massachusetts,^20^ was used for external validation. The analysis included adult patients aged ≥18 years who had at least one blood culture collected during the study period. Comparable data preprocessing methods were applied across both datasets. The study size was determined by the number of eligible patients available in each database during the study period, and all eligible patients were included in the analysis.

### Outcomes and Data Preprocess

We evaluated two prediction tasks using a shared event-based input representation: candidemia within 7 days of the index blood culture and 30-day mortality. Blood cultures were transformed into event-based samples, with the earliest culture defining the index time. A 7-day observation window was assigned to each index event, and candidemia was defined by the presence of any blood culture positive for *Candida* spp. within this window (Figure 1a). Blood cultures obtained more than 7 days after a prior index time initiated a new event.

**Figure 1:**
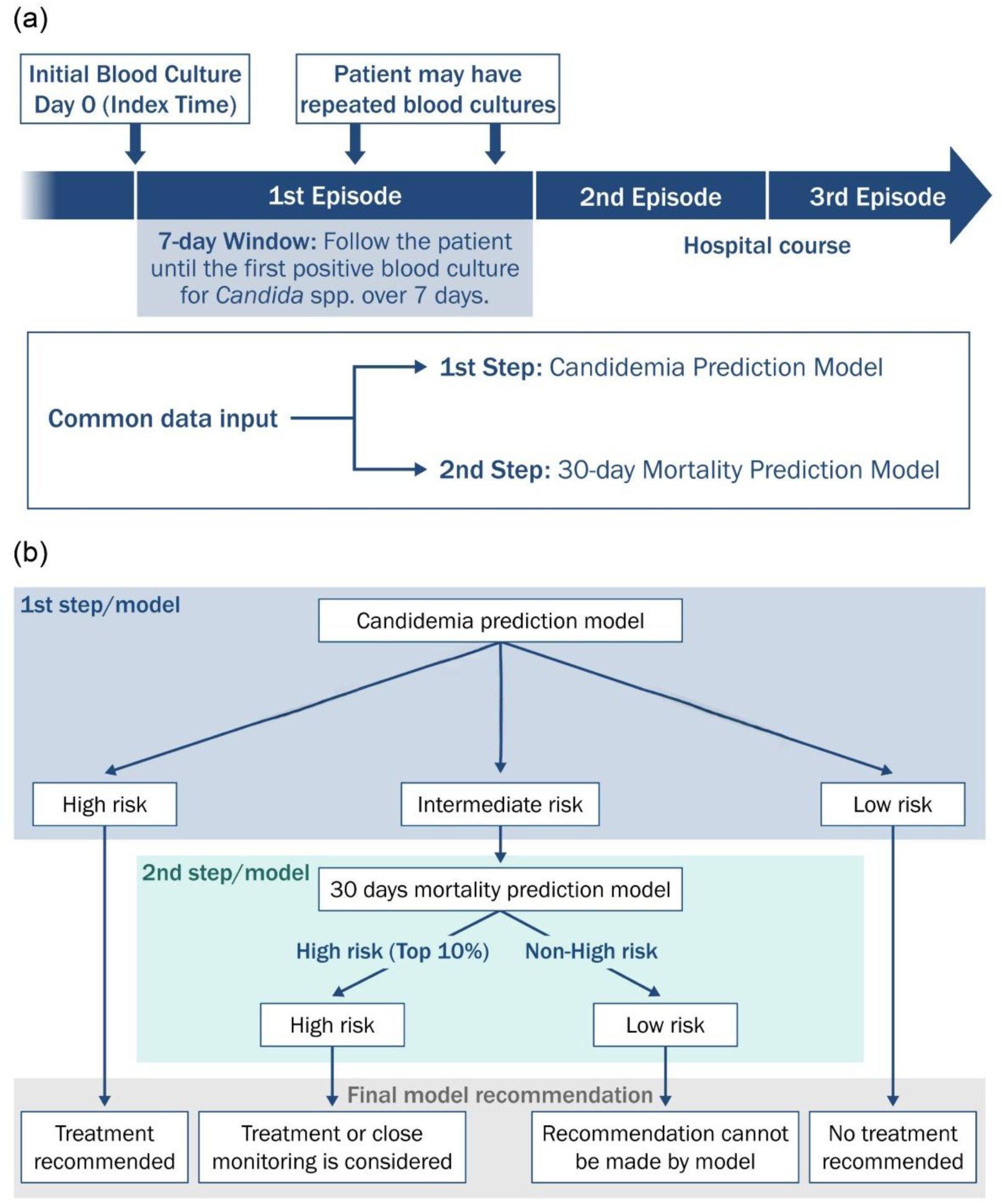
Study design and two-step prediction framework for candidemia risk stratification. (a) Definition of candidemia episodes and index time based on blood culture collection All blood culture events were extracted from each dataset and transformed into event-based samples. For each patient, the earliest blood culture timestamp was defined as the index time (time [t] = 0). A 7-day observation window was assigned to each index time, and candidemia labels (outcome) were generated based on the presence of any blood culture yielding *Candida* spp. within this window. Any blood culture obtained more than 7 days after the preceding index time triggered creation of a new index time and initiation of a new 7-day observation window, thereby forming an additional event. Both predicted outcomes (candidemia and 30-day mortality) were derived using the same event construction and feature extraction pipeline. (b) Two-step prediction framework integrating candidemia and 30-day mortality risk Patients were initially stratified into risk groups using predefined operating thresholds of the candidemia prediction model. The probability threshold corresponding to 95% specificity defined the high-risk group, whereas the threshold corresponding to 90% sensitivity defined the low-risk group; patients falling between these thresholds were classified as intermediate risk. High-risk patients were recommended empiric antifungal therapy, whereas low-risk patients were not. Patients classified as intermediate risk underwent a second-step assessment using a 30-day mortality prediction model. Within this group, individuals in the top 10% of predicted mortality risk were considered high risk and eligible for antifungal treatment. No definitive treatment recommendation was generated for the remaining patients.

For each event, demographic, clinical, laboratory, medication, and hospitalization features were extracted as model inputs (Supplemental Table S1). Temporal features were aggregated at higher resolution near the index time and at progressively coarser resolution for more distant history to preserve clinical acuity while limiting sequence length (details are in supplementary materials). Only data available at the index time were included, without imputation, to avoid label leakage. All variables were encoded as categorical features using the PyTorch_EHR framework. Additional details are provided in the Supplement.

### Model Architecture

We used the PyTorch_EHR framework to train gated recurrent unit (GRU)-based recurrent neural networks on longitudinal EHR data to predict 7-day candidemia and 30-day mortality.^21^ Both tasks shared identical inputs and architecture with task-specific output layers. Categorical events were embedded, and temporal information was incorporated using time-interval encoding and masking to handle irregular sampling. The final hidden state was used to generate outcome probabilities. Additional details are provided in the Supplementary materials.

### Training and Evaluation

Patients were stratified into four groups based on prior candidemia and ICU admission (Supplemental Figure S1) and randomly split into training, validation, and test sets (70%, 10%, and 20%) within each stratum to ensure balanced representation. For 7-day candidemia prediction, the deep learning model was compared with logistic regression (LR)^22^ and LightGBM (LGBM).^23^ Given the low event prevalence, class imbalance was addressed using positive-case oversampling and class-weighted loss. Numerical variables were standardized, and hyperparameters were optimized for all models using Optuna.^24^ Model performance was evaluated using area under the receiver operating characteristic curve (AUROC) and area under the precision–recall curve (AUPRC). For external validation, models trained on HMHS were tested on the independent MIMIC-IV dataset using identical preprocessing and evaluation procedures.

### Feature importance

For LGBM, we used SHapley Additive exPlanations (SHAP) to quantify feature importance.^25^ To interpret the deep learning models, we applied integrated gradients to estimate the contribution of each input feature to the model output.^26^ Feature importance was summarized using the mean absolute integrated gradient across patients.

### Two-Step Prediction Framework

In the first step, patients were stratified into three groups based on candidemia risk—high, intermediate, and low risk. A threshold corresponding to 95% specificity was used to define the high-risk group, for whom empiric antifungal treatment was recommended. This specificity level was selected to minimize false-positive predictions and avoid unnecessary antifungal exposure in a low-prevalence setting. Conversely, patients with predicted risk below the threshold corresponding to 90% sensitivity were classified as low-risk, and antifungal treatment was not recommended. Given the substantial mortality associated with candidemia, a sensitivity threshold of 90% was chosen to capture the majority of cases.

Patients whose predicted risk fell between these two thresholds were categorized as intermediate risk, reflecting uncertainty in the one-step model and necessitating further risk assessment in the second step (Figure 1b). For patients classified as intermediate risk for candidemia, we implemented a second predictive stage using the 30-day mortality model to provide additional prognostic information to guide treatment decisions. In this setting of diagnostic uncertainty, mortality risk may inform the clinical tolerance for potential overtreatment. In this step, patients whose predicted mortality risk fell within the top 10% were classified as high risk, and antifungal treatment was considered clinically appropriate. The top 10% mortality risk was selected to define a clinically meaningful high-risk subgroup while preserving interpretability and adequate group size balance, rather than as a direct treatment threshold. Patients in the remaining 90% were categorized as low risk; for these individuals, the model cannot provide a definitive treatment recommendation. To evaluate the potential clinical impact of our model against prescribers’ decisions, we investigated the use of empirical antifungal therapy within 24 hours among patients in each predicted risk category, stratified by whether candidemia was ultimately confirmed or not.

## Statistical Analysis

Baseline characteristics were summarized using descriptive statistics, and categorical variables were compared between patients with and without candidemia using Fisher’s exact or chi-square tests. Differences in AUROC between models were assessed using the DeLong test. Model AUPRCs were compared using stratified bootstrap resampling (10,000 iterations). Time to candidemia was assessed using 7-day cumulative incidence curves with 95% confidence intervals and compared across risk strata using the log-rank test. Analyses were conducted in IBM SPSS Statistics version 30 and Python version 3.12, with two-sided p-values <0.05 considered significant. The study protocol was approved by the Ethics Committee of the HMHS (Reference ID: PRO00037862). This study is reported in accordance with the Transparent Reporting of a multivariable prediction model for Individual Prognosis Or Diagnosis (TRIPOD) statement (Supplemental Table S2).^27^

## Results

### Patient characteristics

In the HMHS dataset, 213,404 patients had at least one blood culture collected, of whom 851 (0.40%) developed candidemia within 7 days of the index culture. In the MIMIC-IV cohort, 107,507 patients had blood cultures collected, and 634 (0.59%) developed candidemia. Patient characteristics are summarized in Table 1. Across both datasets, candidemia patients had substantially higher rates of major comorbidities, including chronic kidney disease (HMHS: 30.3% vs. 10.1%; MIMIC-IV: 20.5% vs. 5.0%), chronic pulmonary disease (25.1% vs. 13.0%; 25.4% vs. 7.3%), cirrhosis (7.2% vs. 1.9%; 6.5% vs. 0.8%), and diabetes mellitus (27.7% vs. 13.4%; 28.1% vs. 8.6%) (all p<0.001).

**Table 1:**
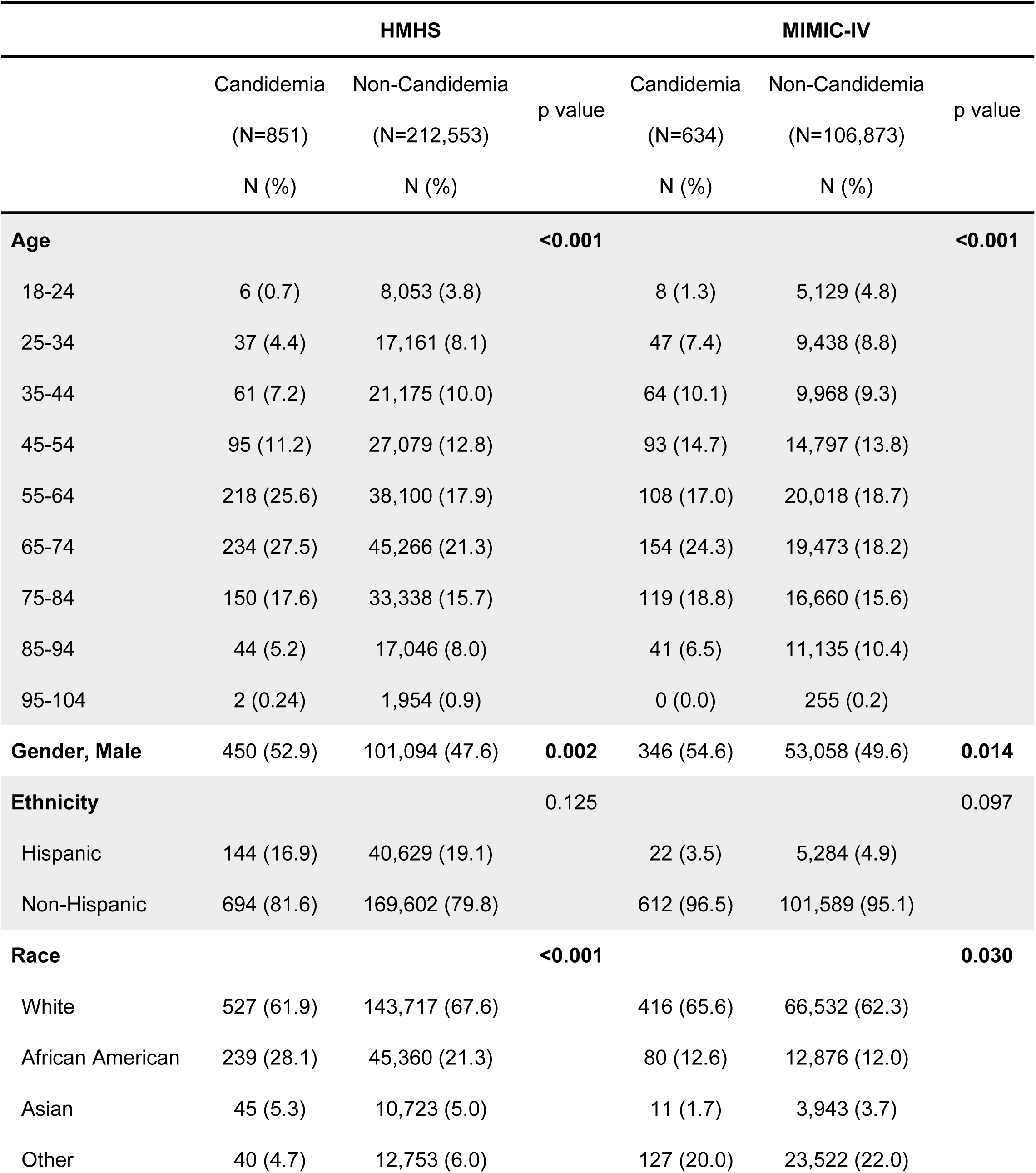
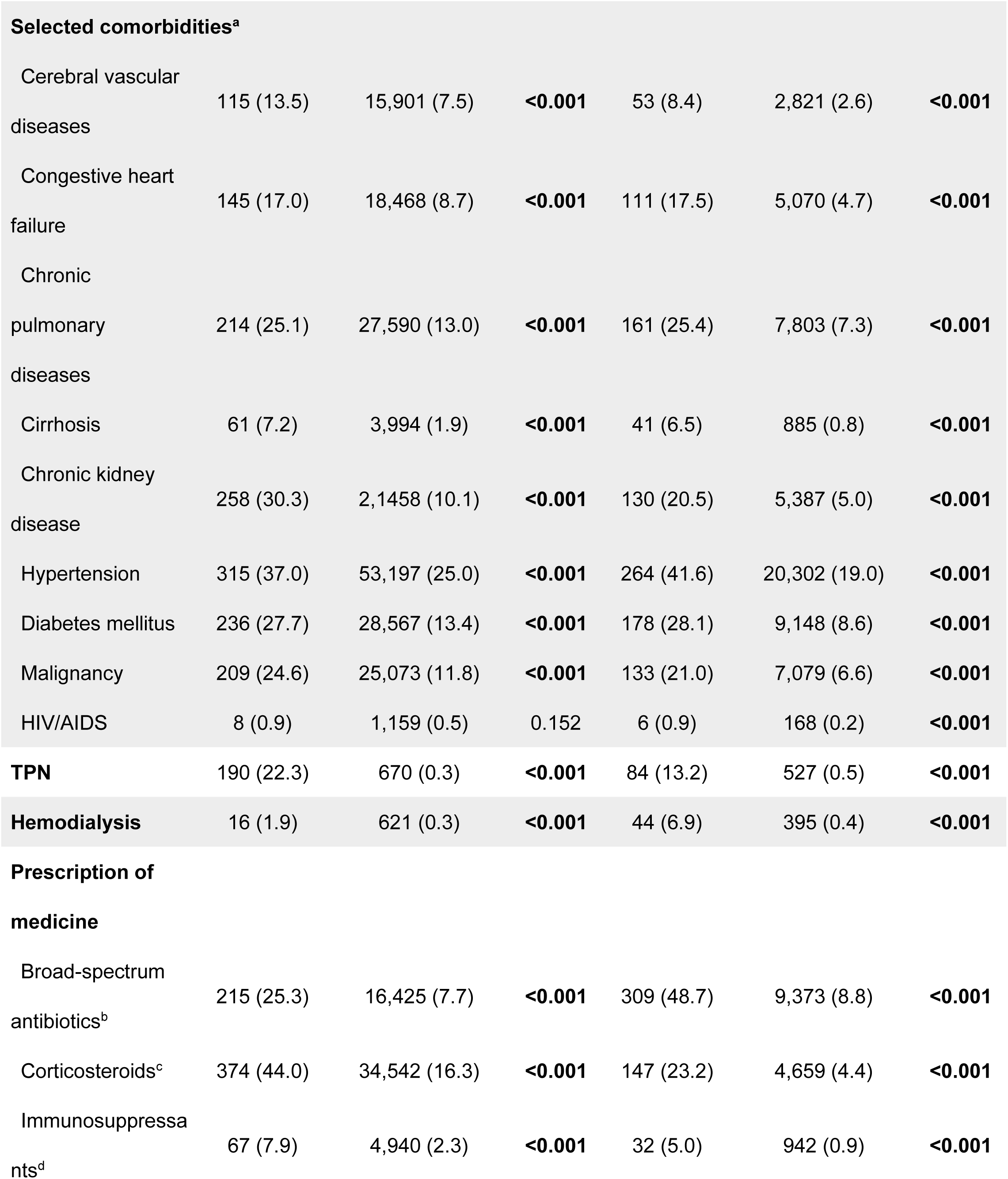
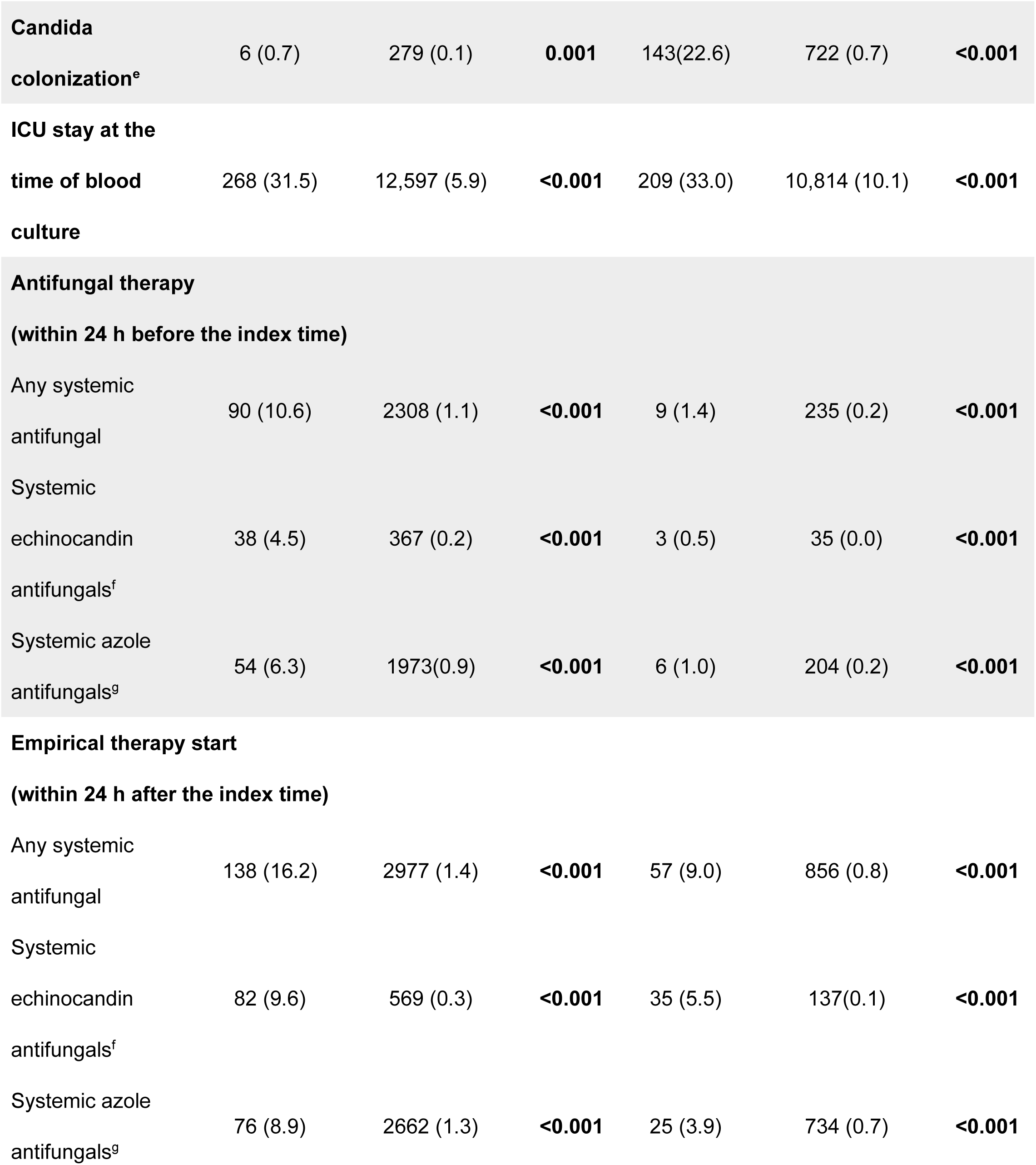
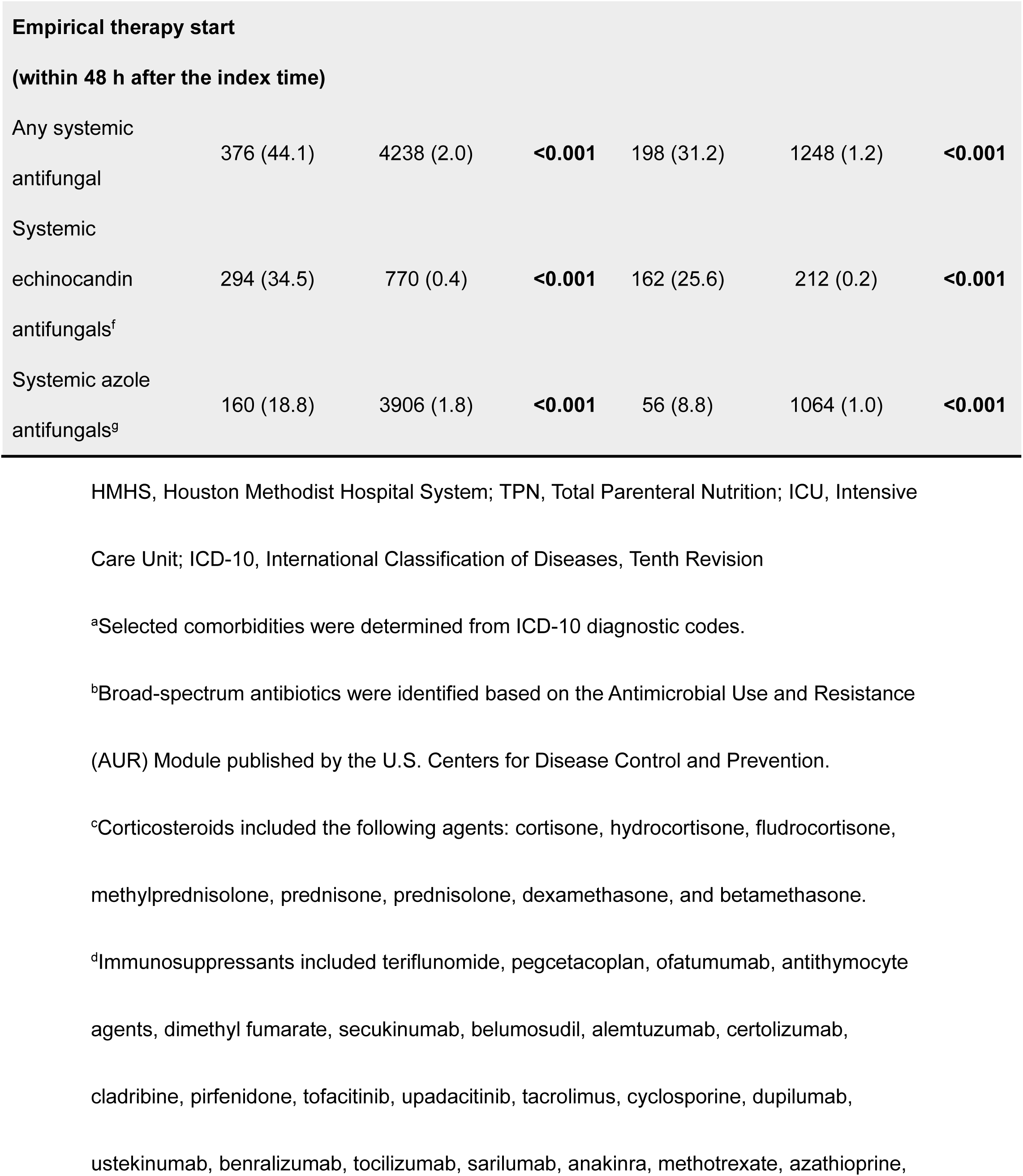
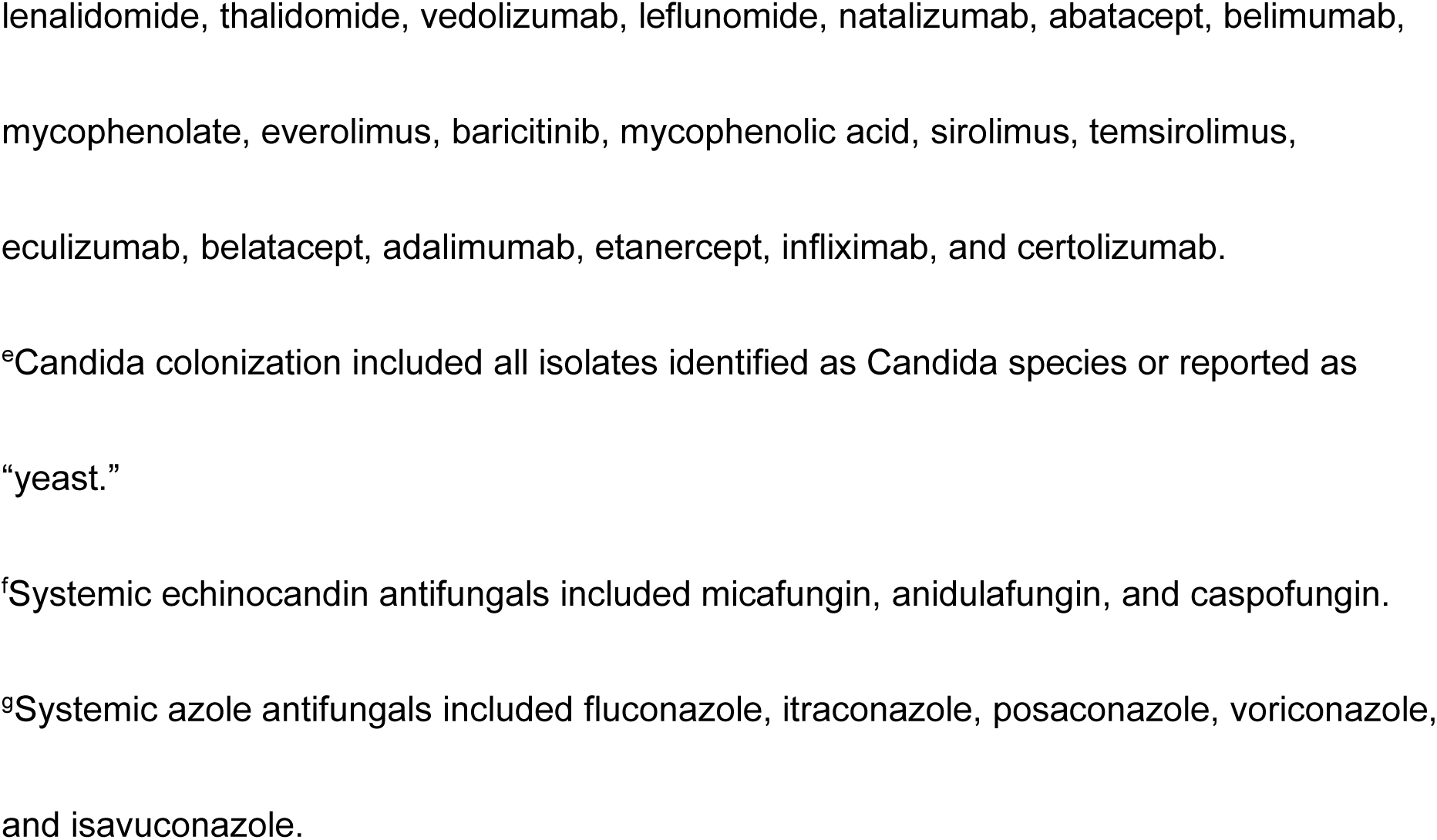
Baseline characteristics of patients with and without candidemia.

Established candidemia risk factors were also more prevalent among cases, including total parenteral nutrition (HMHS: 22.3% vs. 0.3%; MIMIC-IV: 13.2% vs. 0.5%), hemodialysis (1.9% vs. 0.3%; 6.9% vs. 0.4%), broad-spectrum antibiotic exposure (25.3% vs. 7.7%; 48.7% vs. 8.8%), corticosteroid use (44.0% vs. 16.3%; 23.2% vs. 4.4%), and immunosuppressive therapy (7.9% vs. 2.3%; 5.0% vs. 0.9%) (all p<0.001). Documented Candida colonization was more frequent among candidemia cases (HMHS: 0.7% vs. 0.1%; MIMIC-IV: 22.6% vs. 0.7%), as was ICU admission at blood culture collection (HMHS: 31.5% vs. 5.9%; MIMIC-IV: 33.0% vs. 10.1%) (all p<0.001).

Systemic antifungal use was higher among candidemia patients. Within 24 hours before the index culture, antifungal therapy was administered to 10.6% vs. 1.1% of patients in HMHS and 1.4% vs. 0.2% in MIMIC-IV (all p<0.001). Empirical antifungal therapy within 24 hours after the index culture was also more common among candidemia cases (HMHS: 16.2% vs. 1.4%; MIMIC-IV: 9.0% vs. 0.8%). Event-level analyses (Supplemental Table S3) showed similar patterns; however, absolute treatment rates remained low. In HMHS, only 16.4% received empirical therapy, while in MIMIC-IV these proportions were 3.8%.

### Model prediction

Model performance for predicting 7-day candidemia is summarized in Table 2. Across all inpatient wards, PyTorch_EHR demonstrated the best performance in both datasets. In HMHS, it achieved an AUROC of 0.868 and an AUPRC of 0.062, demonstrating the highest AUPRC compared with logistic regression (0.033) and LightGBM (0.036). Similar results were observed in external validation using MIMIC-IV (AUROC 0.764; AUPRC 0.043). Although transfer learning from HMHS to MIMIC-IV resulted in lower performance than retraining on MIMIC-IV, it remained superior to traditional ML models. The length and density of available time-series data differed between the HMHS and MIMIC-IV cohorts (Supplementary Figure S2). Our study demonstrated consistently superior discrimination of PyTorch_EHR across both cohorts, particularly in terms of AUPRC under extreme class imbalance (Supplementary Figure S3).

**Table 2:**
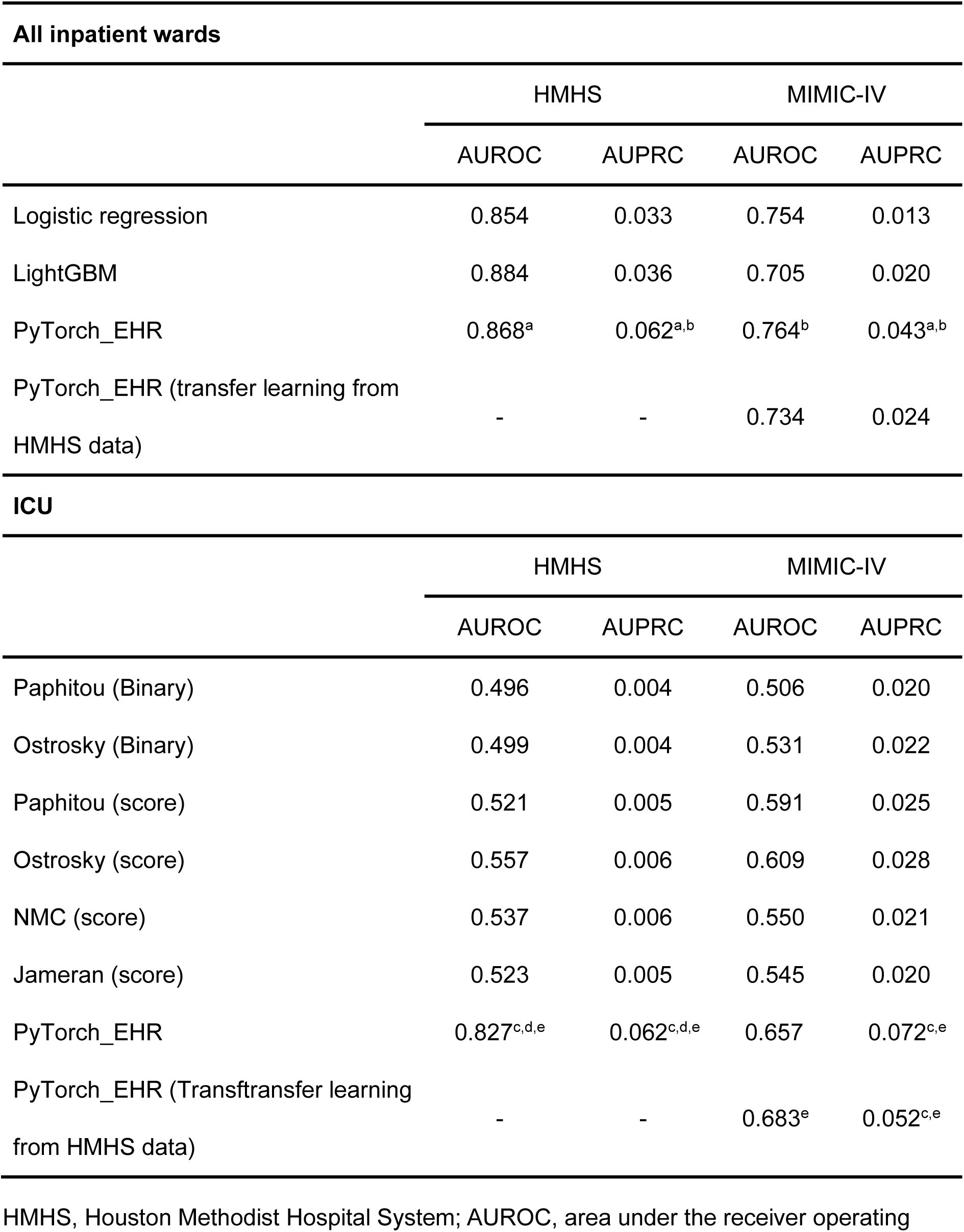
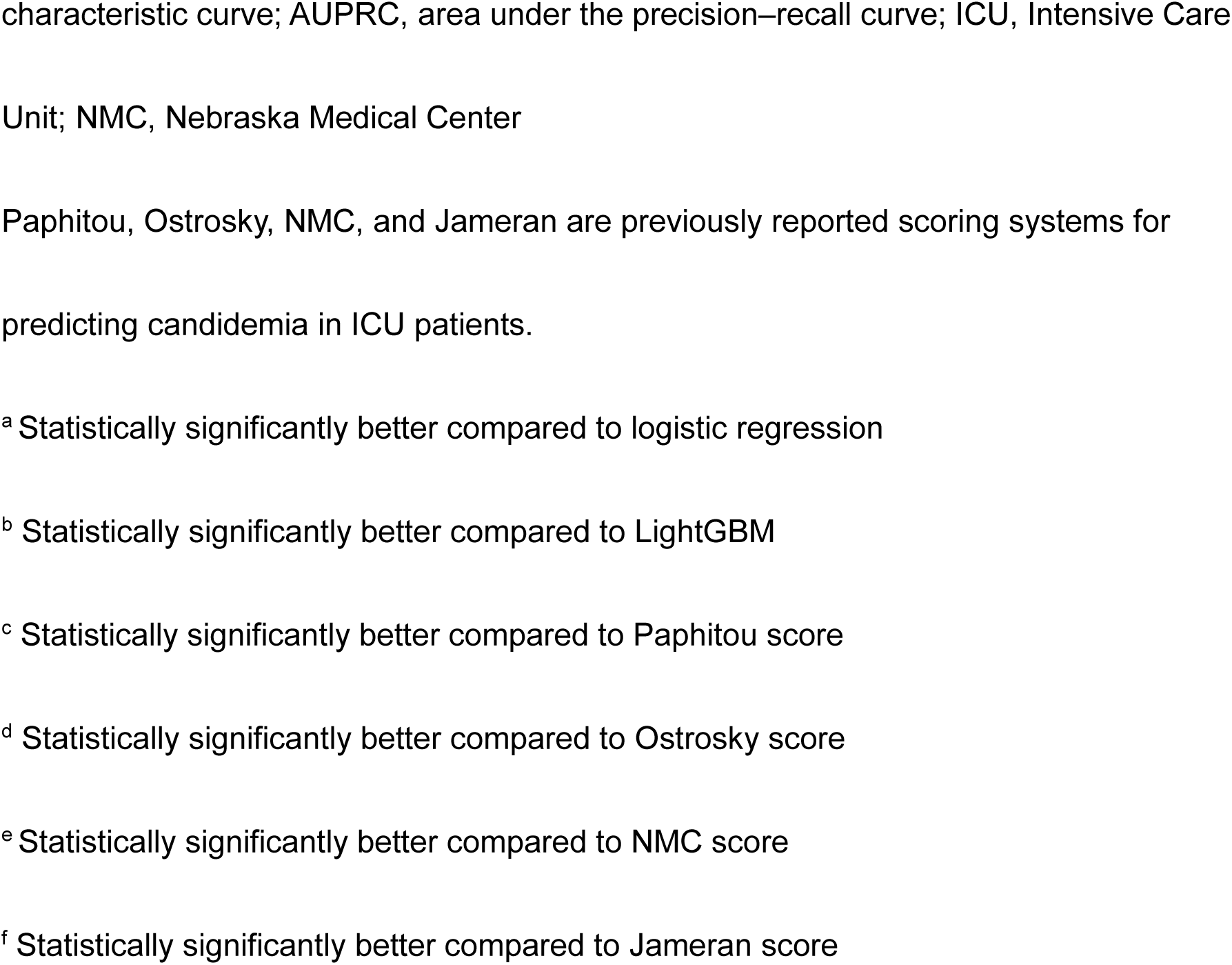
Performance of prediction models for candidemia across all inpatient wards and intensive care units in HMHS and MIMIC-IV.

In ICU subgroups, PyTorch_EHR also outperformed established candidemia scoring systems, including the Paphitou,^11^ Ostrosky,^10^ Nebraska Medical Center,^12^ and Jameran scores.^13^ In the HMHS ICU cohort, PyTorch_EHR achieved an AUROC of 0.827 and an AUPRC of 0.062, with comparable performance in MIMIC-IV (AUROC 0.657; AUPRC 0.072). Transfer learning again resulted in modest performance attenuation (AUROC 0.683; AUPRC 0.052) but remained superior to comparator scores.

Performance of the 30-day mortality model is shown in Supplemental Table S4. PyTorch_EHR demonstrated strong discrimination in both HMHS (AUROC 0.891; AUPRC 0.422) and MIMIC-IV (AUROC 0.826; AUPRC 0.390), with comparable performance under transfer learning.

### Comparison between one-step and two-step prediction models

Supplemental Table S5 presents the diagnostic performance of the candidemia prediction model. Supplemental Table S6 summarizes candidemia case distribution across treatment recommendation categories. In the HMHS test set, the one-step model classified 101 of 199 candidemia cases (50.8%) as treatment recommended. The two-step model reclassified an additional 20 cases, increasing coverage to 121 cases (60.8%). In MIMIC-IV, the one-step model identified 40 of 147 cases (27.2%), while the two-step approach increased coverage to 68 cases (46.2%).

Figure 2 shows 7-day cumulative incidence curves stratified by model-assigned risk groups. Patients in the treatment recommended group consistently had the highest incidence of candidemia. Those classified as intermediate risk for candidemia but high risk for mortality (treatment considered) showed intermediate but clearly elevated risk, while lower-risk groups demonstrated substantially lower incidence. Risk strata were well separated in both cohorts, with log-rank tests confirming significant differences across all groups (all p <0.001).

**Figure 2.**
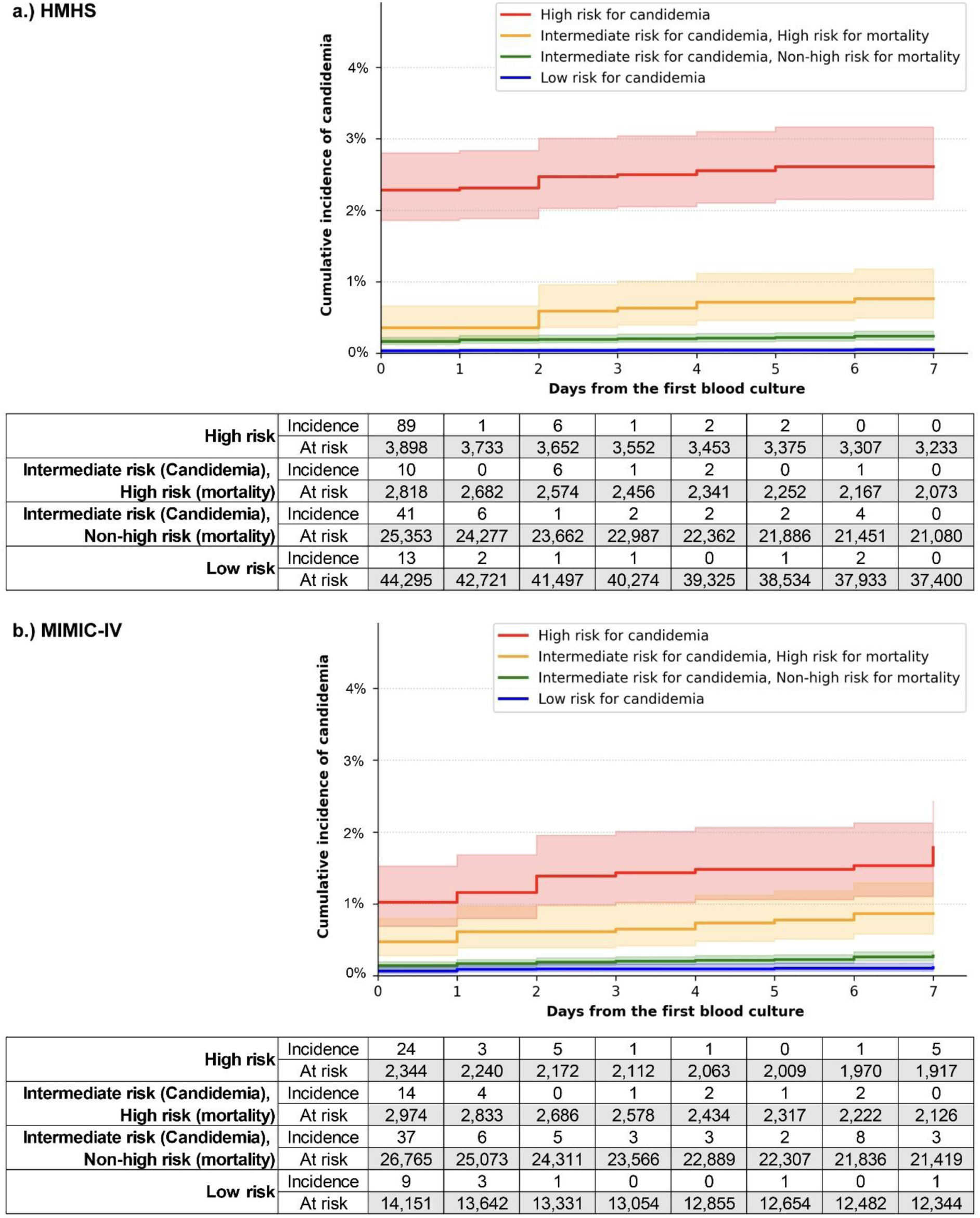
Seven-day cumulative incidence of candidemia stratified by model-assigned risk groups. Seven-day cumulative incidence of candidemia stratified by risk groups defined by the two-step prediction framework in the (a) Houston Methodist Hospital System (HMHS) and (b) MIMIC-IV cohorts. High-risk patients (treatment recommended) showed the highest cumulative incidence, followed by intermediate-risk patients (Treatment or close monitoring is considered). Intermediate-risk patients without a treatment recommendation and low-risk patients (treatment not recommended) showed progressively lower incidence. Shaded areas indicate 95% confidence intervals, and numbers at risk are shown below each panel. Differences among risk groups were significant (log-rank test, all p < 0.001).

### Potential clinical impact of the two-step model on empirical antifungal therapy

Table 3 compares model recommendations with empirical antifungal therapy administered within 24 hours. In HMHS, only 14.9% of candidemia cases classified as treatment recommended received empirical antifungal therapy (corresponding to approximately one true candidemia case per 38 treated patients), while 85.1% did not; among untreated patients, 34.9% died within 30 days. Similarly, 90.0% of patients in the treatment considered group did not receive therapy, and 61.1% of these untreated patients died. Application of the two-step model would enable 104 additional candidemia cases to receive earlier therapy (corresponding to approximately one true candidemia case per 56 treated patients). Comparable findings were observed in MIMIC-IV, 90.0% did not receive empirical antifungal therapy (corresponding to approximately one true candidemia case per 59 treated patients). Additionally, 96.4% of patients in the treatment-considered group were untreated, among whom 74.1% died within 30 days. Use of the two-step framework would allow 63 additional cases to receive earlier therapy (corresponding to approximately one true candidemia case per 78 treated patients. In both datasets, approximately 10% of candidemia cases were classified as low risk and not recommended for treatment.

**Table 3:**
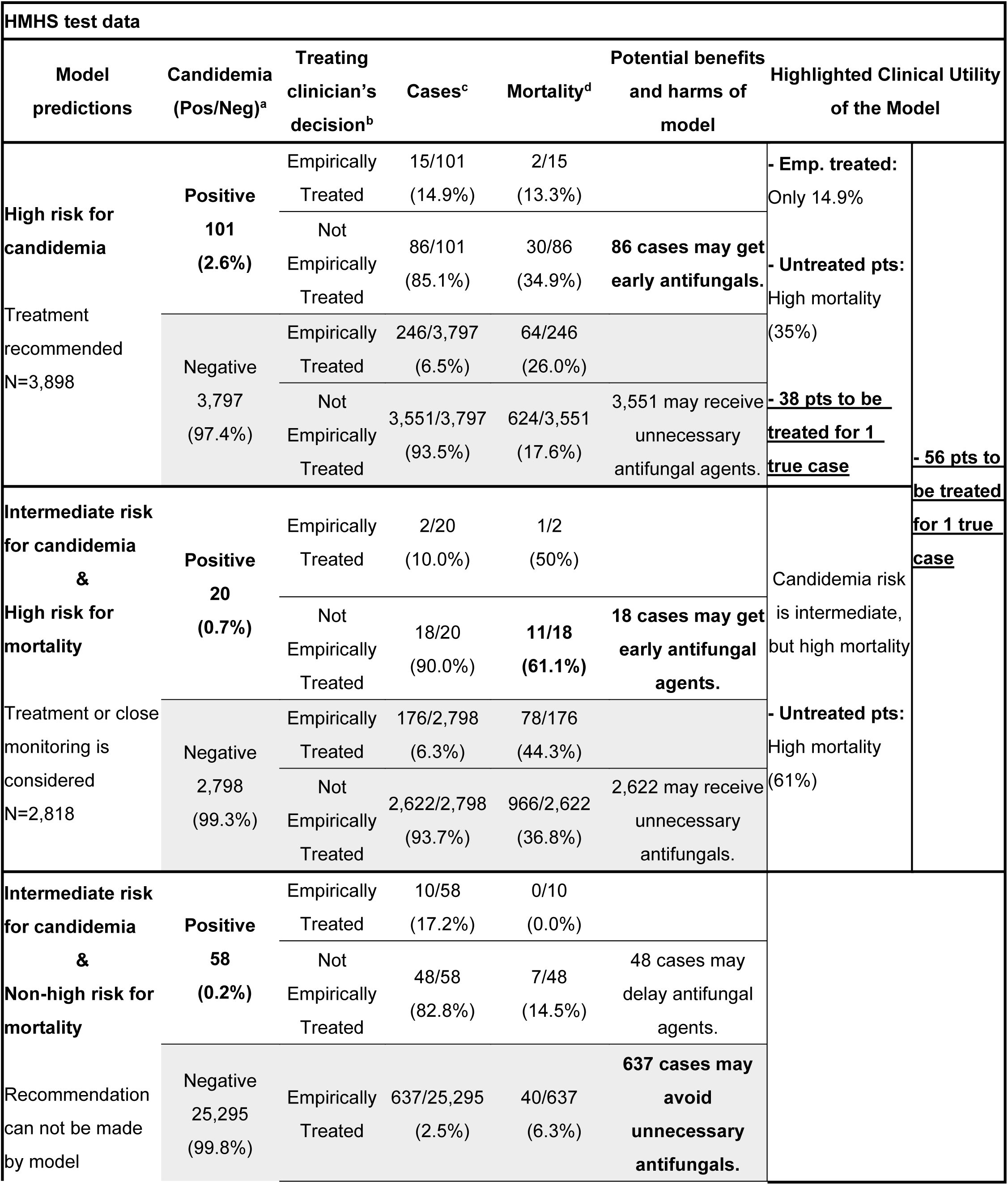
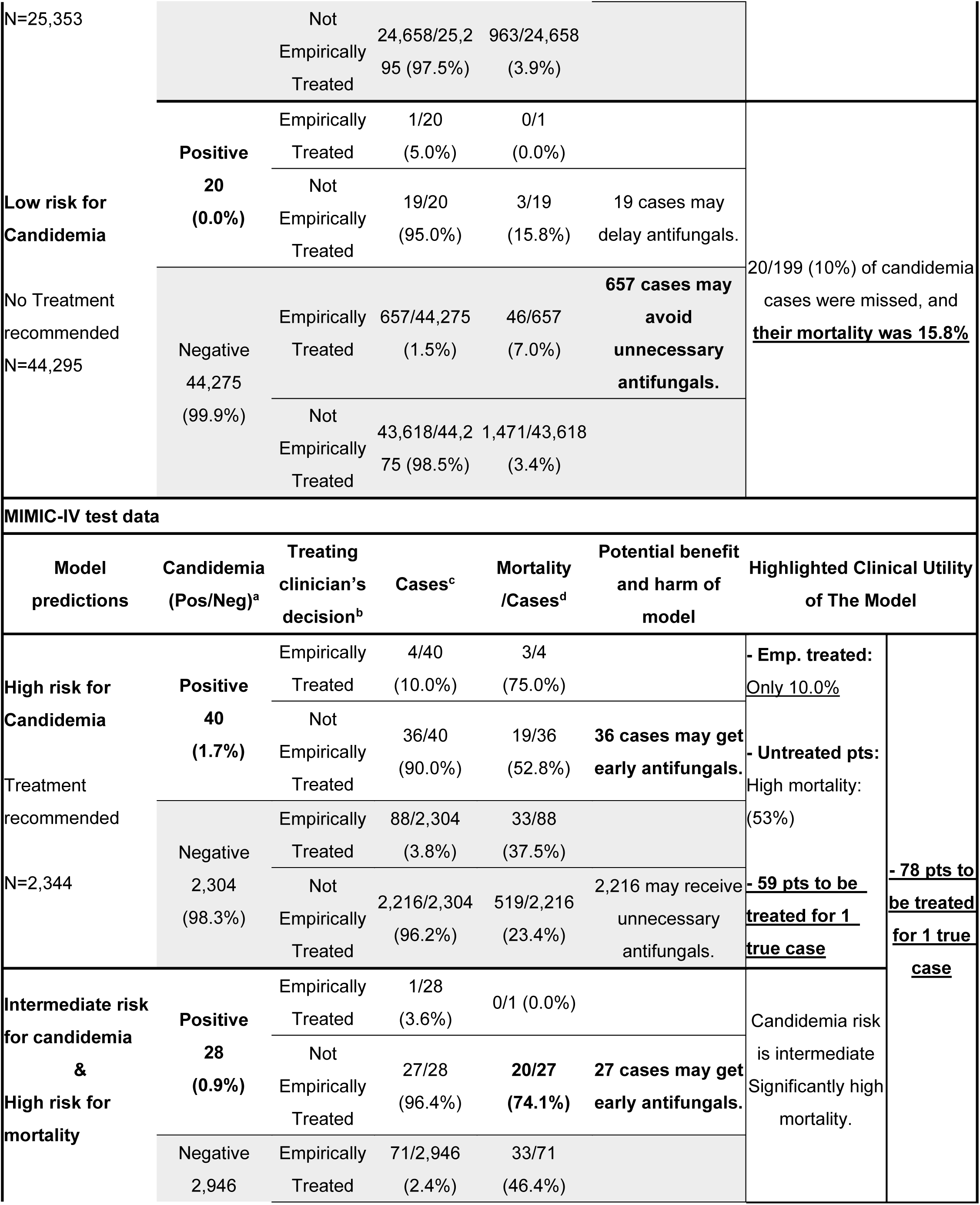
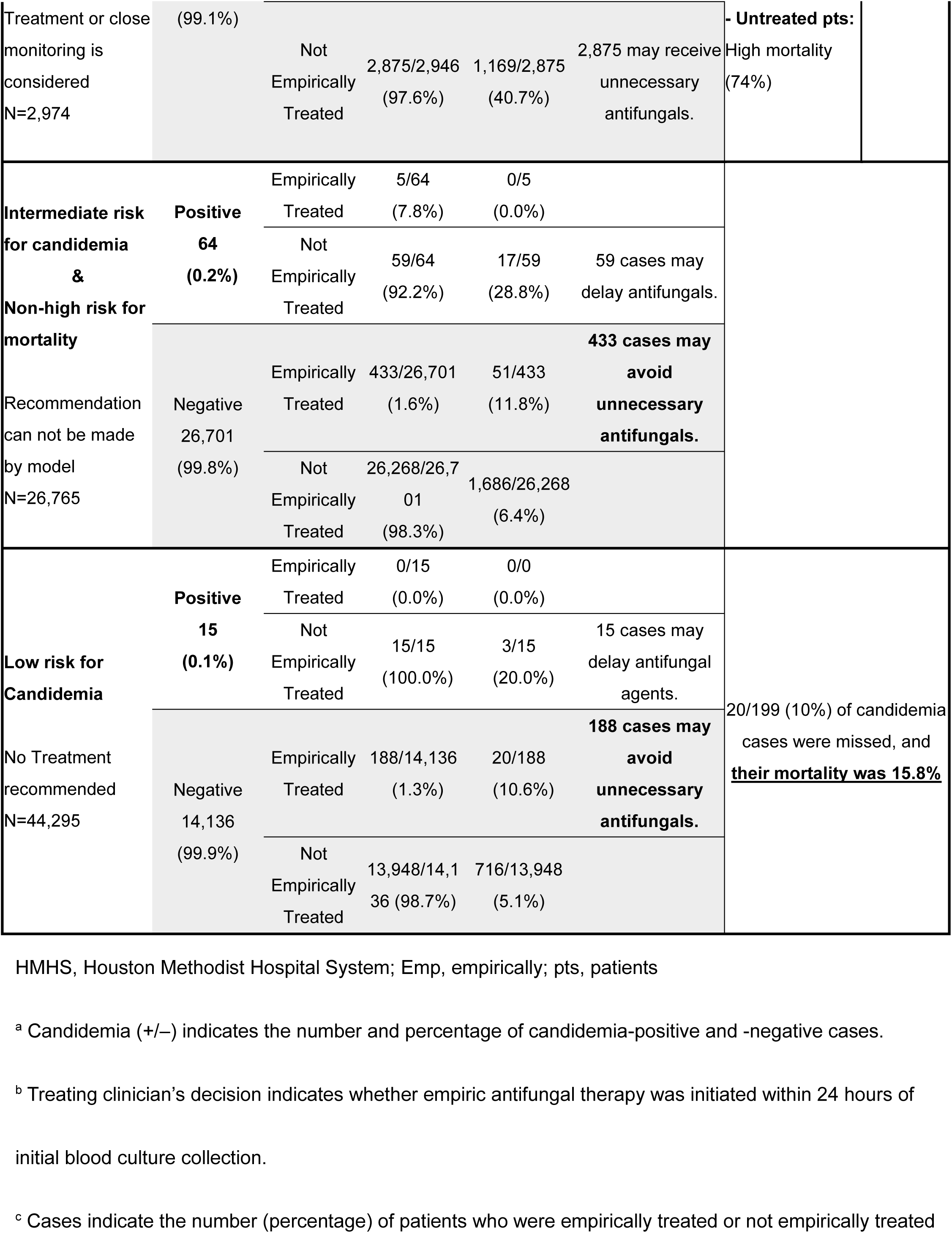
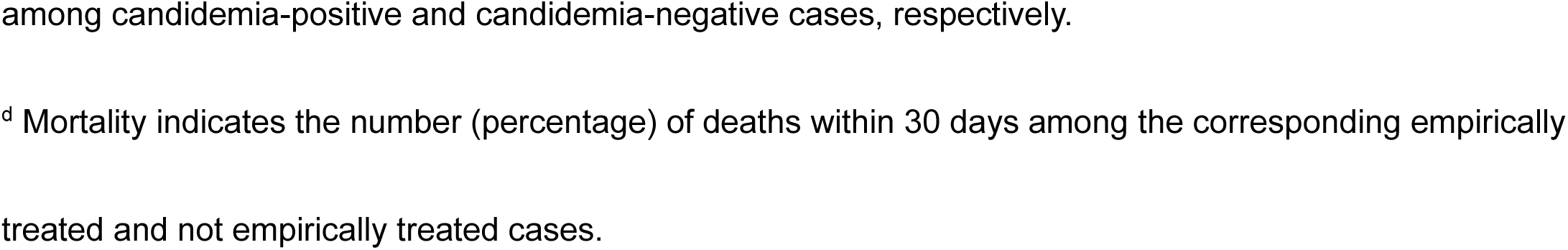
Potential clinical impact of the two-step mod.

### Feature importance

Feature importance for the LightGBM and PyTorch-EHR models is shown in Supplementary Figures S4 and Table S7, respectively. In the PyTorch-EHR model, feature importance more frequently highlighted variables related to blood culture results and known risk factors for candidemia, including severe sepsis, acute kidney failure, and administration of nutritional substances via central or peripheral veins.

## Discussion

Using two large, independent EHR datasets, we demonstrated that a deep learning model based on longitudinal EHR data (PyTorch_EHR) substantially outperformed traditional machine learning models and previously reported ICU candidemia scoring systems. Model performance was consistent across internal testing and external validation. Candidemia incidence was <1% in both the HMHS and MIMIC-IV cohorts, creating an extremely imbalanced setting that complicates model development. To address this challenge, we applied a two-step framework combining candidemia risk and 30-day mortality prediction.

This approach increased capture of candidemia cases for whom clinicians might reasonably consider empiric antifungal therapy, even when candidemia risk alone was insufficient to support treatment decisions. Given the high mortality associated with candidemia, clinicians may accept a lower predictive yield, pursue closer monitoring, repeat blood cultures, or use adjunctive diagnostics to guide management.^28^ By comparing model recommendations with physicians’ empirical antifungal use, we demonstrated that the two-step framework could enable earlier treatment in a substantial number of high-risk untreated patients. Together, these findings support a risk-stratified, two-step strategy integrating candidemia and mortality prediction to improve empirical antifungal decision-making.

Predicting candidemia remains inherently challenging because it is a rare yet clinically critical event. In both the HMHS and MIMIC-IV cohorts, candidemia occurred in only 0.4% and 0.6% of patients at the patient event level, respectively. Even in ICU settings, only 0.6% and 1.2% were positive for candidemia. Under such highly imbalanced conditions, predictive modeling becomes particularly challenging, and this imbalance also has important implications for model evaluation. Conventional discrimination metrics such as AUROC may appear favorable even when a model provides limited clinical value in identifying true positive cases. In contrast, AUPRC more directly reflects performance in rare-event settings and is therefore more appropriate for evaluating candidemia prediction, where the clinical cost of missed cases is substantial.^29^ Previously published prediction models have mainly focused on AUROC when evaluating model performance. In this project, our models were trained on AUPRC and evaluated on both AUPRC and AUROC. Our candidemia prediction model demonstrated significantly superior performance compared with conventional machine learning models, particularly with respect to AUPRC under extreme class imbalance.

A key strength of our approach is explicit modeling of longitudinal, event-based EHR sequences rather than reliance on static data at blood culture collection. Temporal information is particularly important for bloodstream infection prediction. Ming et al. showed that removing time-series information substantially degrades deep learning performance, especially for hospital-onset infections.^30^ By leveraging temporally ordered clinical data—including laboratory trends, procedures, medication exposures, and care transitions—our model captures dynamic trajectories preceding candidemia. We further employed an incremental temporal aggregation strategy, using hourly resolution within 7 days of the index time and daily resolution for events 7–30 days prior. This design preserves clinically meaningful short-term dynamics common in critically ill patients while limiting sequence length and improving computational efficiency. Importantly, it mirrors clinical reasoning, in which recent events are weighed more heavily. Unlike prior candidemia scores developed primarily for ICU populations ^10–13^, our model was designed for all inpatient wards. Even when restricted to ICU patients, it outperformed established scoring systems, supporting applicability across care settings.

Due to severe class imbalance, candidemia-only models inherently generate a large intermediate-risk group for whom model-derived treatment decisions are unclear. These patients are not clinically benign, and reliance on single-step prediction may miss opportunities for early intervention among individuals at high risk of adverse outcomes. The proposed two-step framework addresses this limitation by translating model outputs into clinically interpretable risk categories rather than binary predictions alone. By jointly considering candidemia and mortality risk, the model identifies patients for whom early antifungal therapy may be justified, even when the predictive yield for candidemia is lower.

Beyond predictive accuracy, an important contribution of this study is alignment of model outputs with real-world clinical decision-making. Empiric antifungal therapy is typically initiated in selected critically ill patients with suspected invasive candidiasis based on clinical risk stratification and supportive findings.^31^ Consistent with this practice, many patients without classic high-risk features do not receive antifungal therapy at blood culture collection. Delayed initiation of empiric antifungal therapy has been independently associated with increased mortality in candidemia.^1^ In our study, empiric antifungal therapy within 24 hours was initiated in only ∼16% of candidemia cases in the HMHS cohort and 8% in MIMIC-IV. The two-step framework is designed to support clinicians who may not consider initiating empiric therapy by providing actionable guidance—such as mortality risk—to guide next-step decisions, including treatment initiation or repeat cultures. Compared with observed physician practice, a substantial proportion of model-identified high-risk patients did not receive empiric antifungal therapy despite high short-term mortality. Thus, the two-step approach may complement clinical judgment by identifying overlooked high-risk patients while preserving treatment restraint in low-risk groups.

This study has several limitations. First, although we used two large independent datasets, both analyses were retrospective, and residual confounding or incomplete clinical context cannot be excluded. Prospective evaluation is needed to assess real-time clinical performance. Second, the length and density of available time-series data differed between the HMHS and MIMIC-IV cohorts, reflecting database structure and institutional workflows, which may have attenuated transfer-learning performance. Future studies should incorporate additional institutional datasets beyond publicly available sources. Third, candidemia was defined by blood culture positivity, an imperfect reference standard with limited sensitivity ^32^. Although we evaluated β-D-glucan and molecular diagnostics as surrogate markers, inconsistent availability precluded their use as outcome definitions. Fourth, time-sequenced deep learning models are inherently difficult to interpret; we therefore provided summary feature importance estimates (Supplemental table S7). Finally, despite promising results, overall performance remains limited, suggesting that further improvements may require multimodal integration of additional data sources, such as molecular diagnostics.

## Conclusions

Candidemia is a rare yet high-impact clinical event that is difficult to predict using conventional approaches. By leveraging longitudinal EHR data, our deep learning model achieved robust performance across two independent cohorts. The proposed two-step framework further addresses a key limitation of single-step prediction by translating risk estimates into clinically actionable treatment categories. By aligning model outputs with real-world physician decision-making, this approach has the potential to support candidemia management by enabling earlier identification of not only high-risk but also intermediate-risk patients with high-mortality. Prospective studies are warranted to validate the findings and evaluate real-time implementation and clinical impact.

## Author Contributions

Hisato Yoshida, Max W. Adelman, and Masayuki Nigo developed the study concept and design. Hisato Yoshida, María Alejandra Pérez, Francis Ifiora, Francisco Guerra Jr, and Masayuki Nigo collected and analyzed the data. Hisato Yoshida, Max W. Adelman, Hitoshi Yoshimura, Cesar A. Arias, and Masayuki Nigo contributed to the interpretation of the results. Hisato Yoshida, Laila Rasmy, Ziqian Xie, Degui Zhi, Masayuki Nigo conducted interpretation of deep learning methods and results. Stephen L. Jones established the environment required to access and use the dataset. All authors had access to and verified the underlying data. All authors contributed to writing – review & editing, critically reviewed the manuscript, and approved the final version for publication.

## Data availability

HMHS data that supports the findings of this study are not openly available due to reasons of sensitivity and are available from the corresponding author upon requests and our institutional IRB approvals. MIMIC-IV data v3.1 is publicly available after data use agreement on the website. (https://physionet.org/content/mimiciv/3.1/)

## Data Availability

The HMHS data are not publicly available due to institutional data use restrictions. MIMIC-IV data are available through PhysioNet upon completion of required training and data use agreements.

https://physionet.org/content/mimiciv/3.1/

## Acknowledgments

We thank Ms. Joanne Park for helping with the graphic preparation.

