## Supplementary Appendix for "Two-step deep-learning candidemia prediction model using two large time-sequence electronic health datasets"

### Supplementary Materials

**Contents**

**Supplementary Table S1:** Features extracted from datasets

**Supplementary Methods**

**Supplementary Figure S1:** Data splitting strategy

**Supplementary Table S2:** TRIPOD checklist for prediction model validation

**Supplementary Table S3:** Event-level use of systemic antifungal therapy before and after the index blood culture in the HMHS and MIMIC-IV cohorts

**Supplementary Table S4:** Diagnostic performance of the candidemia prediction model in the HMHS and MIMIC-IV cohorts

**Supplementary Figure S2:** Differences in time-series data length and density between the HMHS and MIMIC-IV cohorts

**Supplementary Figure S3:** Receiver operating characteristic and precision–recall curves for candidemia prediction models in the HMHS and MIMIC-IV cohorts

**Supplementary Table S5:** Model performance for 30 days mortality prediction model (HMHS and MIMIC-IV)

**Supplemental Table S6:** Distribution of candidemia cases across treatment recommendation categories for one-step and two-step models

**Supplementary Figure S4:** Feature importance of the LightGBM models for candidemia prediction in the HMHS and MIMIC-IV cohorts.

**Supplemental Table S7:** Feature importance derived from integrated gradients for PyTorch-EHR candidemia prediction models in the HMHS and MIMIC-IV cohorts

**Supplementary Table S1:** Features extracted from datasets

| |
|---|
| Demographic |
| Age (exclude <18 years old) |
| Gender |
| Race |
| Ethnicity |
| Preferred language[a] |
| Admission |
| Hospital admission date, Hospital discharge date, Length of previous hospital stay |
| ICU |
| ICU admission time, ICU discharge time, Length of previous ICU stay |
| Vital signs[b] |
| Laboratory orders and results: LOINC code |
| CPT code |
| ICD-10 code (diagnosis) |
| ICD-10 code (procedure) |
| Microorganism |
| Antibiotic susceptibility, Culture type, *Candida* colonization |
| Medication administration (inpatient) and prescriptions (outpatient): NDC code |
| TPN use |
| Mechanical Ventilator usage and the settings |
| Hemodialysis and continuous renal replacement therapy |

ICU, Intensive Care Unit; LOINC, Logical Observation Identifiers Names and Codes; CPT, Current Procedural Terminology; ICD-10, International Classification of Diseases, Tenth Revision; NDC, National Drug Code; TPN, Total Parenteral Nutrition

[a] Preferred language was available in the Houston Methodist dataset but not in the MIMIC dataset.

[b]Vital signs were recorded only for ICU patients in MIMIC-IV.

### Supplemental Methods

#### Data Preprocess Details

The study evaluated two prediction tasks using the same input features and event-based samples. The primary outcome was candidemia within 7 days of the index blood culture (day zero). The secondary outcome was 30-day mortality, defined as death occurring within 30 days after the index time for each event. Both predicted outcomes were derived using the same event construction and feature extraction pipeline.

All blood culture events were extracted from each dataset and transformed into event-based samples. For each patient, the earliest blood culture timestamp was defined as the index time (time [t] = 0). A 7-day observation window was assigned to each index time, and candidemia labels (outcome) were generated based on the presence of any blood culture yielding *Candida* spp. within this window (Main manuscript Figure 1a). When multiple blood cultures were obtained on the same calendar date, they were collapsed into a single event represented by the earliest timestamp. If any culture collected on that date was positive for *Candida*, the event was labeled positive. Blood cultures collected within the same 7-day window were treated as part of a single sampling episode. Any blood culture obtained more than 7 days after the preceding index time triggered creation of a new index time and initiation of a new 7-day observation window, thereby forming an additional event.

For each event, clinical features—summarized in the Supplemental Table 1, including demographics, laboratory measurements, medication administrations, and hospitalization information—were extracted from the period preceding the index time and used as model inputs. Temporal features of inputs were aggregated at varying levels of granularity depending on their proximity to the index time: events within 0–7 days were aggregated at an hourly resolution, those 7–30 days prior at a daily resolution, and those 30–90 days prior at a monthly resolution. Events more than 90 days before the index time were aggregated without time-step segmentation and treated as long-term historical information. This approach allows to provide granular inputs closer to index time to reflect the patient acuity while avoiding a long sequence of events. All input variables were converted into categorical features. To avoid label leakage, only data available at the index time were included. Missing events were not imputed. For example, in the MIMIC-IV dataset, some laboratory tests lacked associated LOINC codes and certain medication records lacked NDC identifiers. These missing entries were left as empty events and represented using masking vectors, allowing the model to appropriately interpret sparse or incomplete temporal sequences. All categorical variables were converted into integer indices following the PyTorch_EHR encoding schema to enable embedding-based representation within the model. Laboratory measurements were transformed into categorical indicators (e.g., high, low, or normal ranges), rather than raw continuous values, allowing them to be treated as discrete event types within the PyTorch_EHR framework. The resulting features were organized into a three-dimensional tensor (events × time steps × feature dimension) accompanied by a parallel masking tensor.

**Model Architecture**

We used the deep learning platform PyTorch_EHR[1] to predict clinical outcomes from event-based EHR sequences. Two prediction tasks were constructed: (1) candidemia within 7 days of the index blood culture, and (2) 30-day mortality. Both tasks used the same input representation but were trained with separate output layers. PyTorch_EHR implements a recurrent neural network (RNN) architecture for modeling longitudinal clinical data. Specifically, we used a gated recurrent unit (GRU) model to model irregular and sparse clinical time-series data, consistent with prior studies demonstrating the effectiveness of recurrent neural networks for EHR-based prediction.[2] Categorical features were mapped to learnable embedding vectors, and temporal information was incorporated using the masking mechanism and time-interval encoding built into the PyTorch_EHR framework. Each event sequence was structured as a time-ordered tensor accompanied by a corresponding masking tensor to handle missing or irregularly spaced events. The GRU processed these sequential representations, and the final hidden state was passed through fully connected layers to generate outcome probabilities. The model source code is publicly available, enabling reproducibility and application in other clinical prediction tasks.[3] For our clinical setting, the interval between events was expressed as the time difference (in days or hours) from the index time, allowing the model to capture acute temporal patterns relevant for early candidemia detection and mortality risk stratification.

**Training and Evaluation**

We first divided all patients into four groups based on their history of candidemia and ICU admission (Supplemental Figure 1). Each group was then randomly split into training, validation, and test sets in a 70%, 10%, and 20% ratio. This stratified approach ensured that both ICU and non-ICU patients, as well as those with and without prior candidemia, were evenly represented across all datasets. When multiple events were available for a single patient, all events from that patient were assigned to the same dataset to prevent information leakage across training, validation, and test sets.

For internal validation, predicted risks were calculated by applying the trained model to the validation subset using identical predictor definitions and preprocessing procedures. For external validation, predictions were generated by applying the fixed model trained on the HMHS cohort to the MIMIC-IV dataset without model retraining. No model updating or recalibration was performed following validation. The MIMIC-IV cohort differed from the development cohort (HMHS) in data source and institutional setting, while eligibility criteria, outcome definitions, and predictor representations were otherwise consistent across cohorts, although variable availability and coding systems differed between databases. Unadjusted associations between individual predictors and outcomes were not assessed.

For the binary classification task of predicting candidemia within 7 days of the index blood culture, we compared the performance of our deep learning model with two traditional machine learning algorithms: logistic regression (LR)[4] and LightGBM (LGBM).[5]

Given the extremely low prevalence of candidemia within 7 days in both HMHS and MIMIC-IV, we applied class-balancing techniques—specifically oversampling of positive cases and class-weighted loss—to address the severe class imbalance during model training.

Before model training, numerical variables were standardized to improve the performance of traditional machine learning algorithms. Hyperparameter tuning for all models, including the RNN-based approach, was performed using Optuna.[6] Model performance was assessed using the area under the receiver operating characteristic curve (AUC) and the area under the precision–recall curve (AUPRC). To evaluate generalizability, the final models trained on the HMHS dataset were externally validated using the independent MIMIC-IV test set, following the same preprocessing, input structure, and evaluation procedures.

**Supplementary Figure S1: Data splitting strategy**

The dataset was split into training, validation, and test sets at a ratio of 70:10:20. Because individual patients could contribute multiple longitudinal events, dataset partitioning was performed at the patient level using unique patient identifiers to prevent information leakage across splits. Given the rarity of candidemia and the strong association between intensive care unit (ICU) exposure and candidemia risk, stratified sampling was applied to ensure balanced distributions of candidemia cases and ICU admissions across the training, validation, and test datasets.

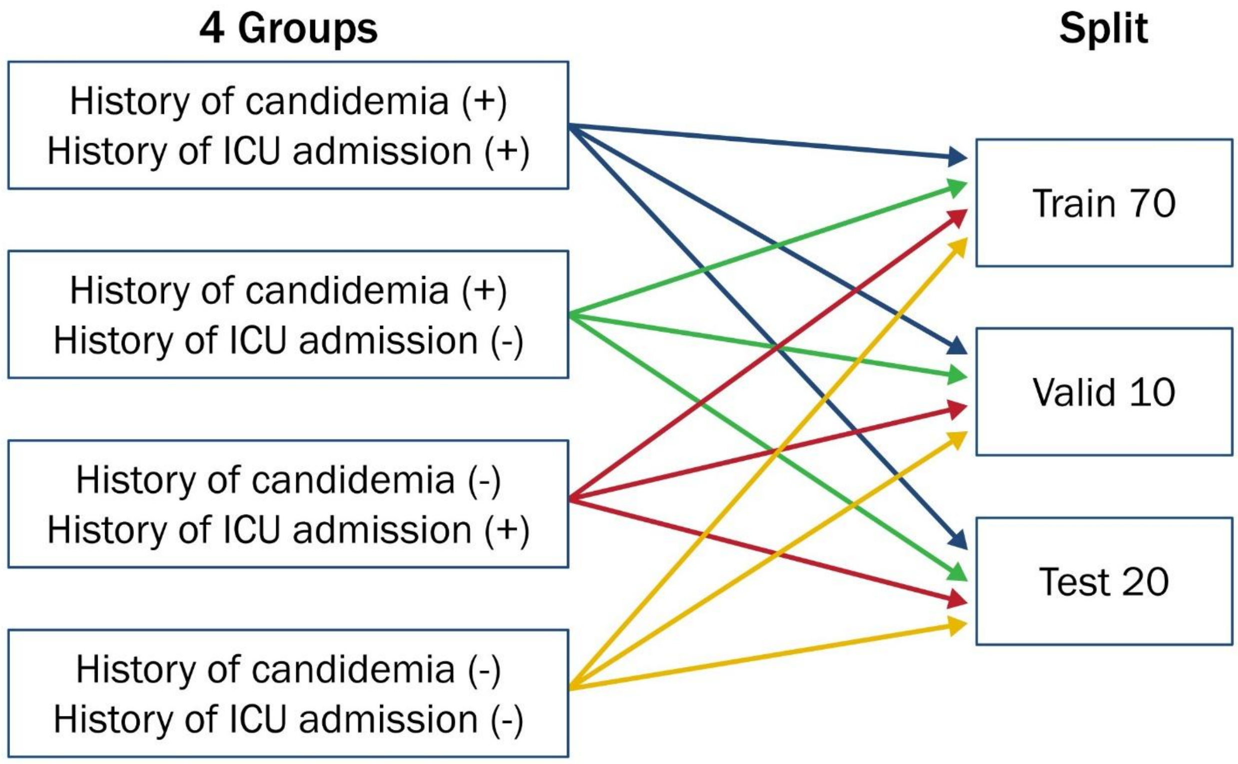

**Supplementary Table S2: TRIPOD checklist for prediction model validation**

| Section/Topic | Item | Section/Topic Item Development | Page |
|---|---|---|---|
| **Title and abstract** | | | |
| Title | 1 | Identify the study as developing and/or validating a multivariable prediction model, the target population, and the outcome to be predicted | 1 |
| Abstract | 2 | Provide a summary of objectives, study design, setting, participants, sample size, predictors, outcome, statistical analysis, results, and conclusions | 3 |
| **Introduction** | | | |
| Background and objectives | 3a | Explain the medical context (including whether diagnostic or prognostic) and rationale for developing or validating the multivariable prediction model, including references to existing models | 5-7 |
| Background and objectives | 3b | Specify the objectives, including whether the study describes the development or validation of the model or both | 5-7 |
| **Methods** | | | |
| Source of data | 4a | Describe the study design or source of data (e.g., randomized trial, cohort, or registry data), separately for the development and validation data sets, if applicable | 7 |
| Source of data | 4b | Specify the key study dates, including start of accrual; end of accrual; and, if applicable, end of follow-up | 7 |
| Participants | 5a | Specify key elements of the study setting (e.g., primary care, secondary care, general population) including number and location of centres | 7 |
| Participants | 5b | Describe eligibility criteria for participants | 7-8 |
| Participants | 5c | Give details of treatments received, if relevant | 7-8 |
| Outcome | 6a | Clearly define the outcome that is predicted by the prediction model, including how and when assessed | 8 |
| Outcome | 6b | Report any actions to blind assessment of the outcome to be predicted | N/A |
| Predictors | 7a | Clearly define all predictors used in developing the multivariable prediction model, including how and when they were measured | 8 |
| Predictors | 7b | Report any actions to blind assessment of predictors for the outcome and other predictors | N/A |
| Sample size | 8 | Explain how the study size was arrived at | 7 |
| Missing data | 9 | Describe how missing data were handled (e.g., complete-case analysis, single imputation, multiple imputation) with details of any imputation method | Supp |
| Statistical analysis methods | 10a | Describe how predictors were handled in the analyses | 8, Supp |

| | | | |
|---|---|---|---|
| Statistical analysis methods | 10b | Specify type of model, all model-building procedures (including any predictor selection), and method for internal validation | 9 |
| Statistical analysis methods | 10c | For validation, describe how predictions were calculated | Supp |
| Statistical analysis methods | 10d | Specify all measures used to assess model performance and, if relevant, to compare multiple models | 9-11, Supp |
| Statistical analysis methods | 10e | Describe any model updating (e.g., recalibration) arising from the validation, if done | Supp |
| Risk groups | 11 | Provide details on how risk groups were created | 10-11 |
| Development vs validation | 12 | For validation, identify any differences from the development data in setting, eligibility criteria, outcome, and predictors | Supp |
| **Results** | | | |
| Participants | 13a | Describe the flow of participants through the study, including the number of participants with and without the outcome and, if applicable, a summary of the follow-up time. A diagram may be helpful | 12 |
| Participants | 13b | Describe the characteristics of the participants (basic demographics, clinical features, available predictors), including the number of participants with missing data for predictors and outcome | 12,13 |
| Participants | 13c | For validation, show a comparison with the development data of the distribution of important variables (demographics, predictors and outcome) | 12,13 |
| Model development | 14a | Specify the number of participants and outcome events in each analysis | 12,13, Table 1 |
| Model development | 14b | If done, report the unadjusted association between each candidate predictor and outcome | Table 1 |
| Model specification | 15a | Present the full prediction model to allow predictions for individuals (i.e., all regression coefficients, and model intercept or baseline survival at a given time point) | N/A[a] |
| Model specification | 15b | Explain how to use the prediction model | 15,16 |
| Model performance | 16 | Report performance with CIs | N/A[b] |
| Model updating | 17 | If done, report the results from any model updating (i.e., model specification, model performance) | N/A[c] |
| **Discussion** | | | |
| Limitations | 18 | Discuss any limitations of the study (such as nonrepresentative sample, few events per predictor, missing data) | 21 |
| Interpretation | 19a | For validation, discuss the results with reference to performance in the development data, and any other validation data | 17-20 |

| | | | |
|---|---|---|---|
| Interpretation | 19b | Give an overall interpretation of the results, considering objectives, limitations, results from similar studies, and other relevant evidence | 21 |
| Implications | 20 | Discuss the potential clinical use of the model and implications for future research | 17-21 |
| **Other information** | | | |
| Supplementary info | 21 | Provide information about the availability of supplementary resources, such as study protocol, Web calculator, and data sets | 23 |
| Funding | 22 | Give the source of funding and the role of the funders for the present study | 23 |

TRIPOD, Transparent Reporting of a multivariable prediction model for Individual Prognosis Or Diagnosis[7]

[a] Not applicable, as a deep learning–based model was used and regression coefficients are not applicable.

[b] Model performance was compared using DeLong's test for AUROC and bootstrap-based hypothesis testing for AUPRC. Confidence intervals were not reported, as the primary focus was on statistical comparison between models.

[c] Model updating was not performed.

**Supplementary Table S3: Event-level use of systemic antifungal therapy before and after the index blood culture in the HMHS and MIMIC-IV cohorts**

| | HMHS | | MIMIC-IV | |
|---|---|---|---|---|
| | Candidemia (N=993) N (%) | Non-Candidemia (N=379,968) N (%) | Candidemia (N=748) N (%) | Non-Candidemia (N=231,650) N (%) |
| **Antifungal therapy (within 24 h before the index time)** | | | | |
| Any systemic antifungal | 134 (13.5) | 12,255 (3.2) | 25 (3.3) | 1,576 (0.7) |
| Systemic echinocandin antifungals[a] | 75 (7.6) | 2,361 (0.6) | 12 (1.6) | 312 (0.1) |
| Systemic azole antifungals[b] | 65 (6.5) | 10,240 (2.7) | 14 (1.8) | 1,289 (0.6) |
| **Empirical therapy start (within 24 h after the index time)** | | | | |
| Any systemic antifungal | 163 (16.4) | 8,635 (2.3) | 29 (3.8) | 2,148 (0.9) |
| Systemic echinocandin antifungals[a] | 104 (10.5) | 2,431 (0.6) | 20 (2.6) | 240 (0.1) |
| Systemic azole antifungals[b] | 75 (7.6) | 7,126 (1.9) | 11 (1.5) | 1,940 (0.8) |
| **Empirical therapy start (within 48 h after the index time)** | | | | |
| Any systemic antifungal | 431 (43.4) | 11,969 (3.2) | 85 (11.4) | 2,863 (1.2) |
| Systemic echinocandin antifungals[a] | 339 (34.3) | 3,308 (0.8) | 72 (9.6) | 367 (0.2) |
| Systemic azole antifungals[b] | 179 (18.0) | 10,204 (2.7) | 24 (3.2) | 2,575 (1.1) |

HMHS, Houston Methodist Hospital System

[a]Systemic echinocandin antifungals included micafungin, anidulafungin, and caspofungin.

[b]Systemic azole antifungals included fluconazole, itraconazole, posaconazole, voriconazole, and isavuconazole.

**Supplementary Figure S2: Differences in time-series data length and density between the HMHS and MIMIC-IV cohorts**

This figure illustrates differences in the length and density of available time-series data between the HMHS and MIMIC-IV cohorts. Panels (a) and (b) show the distribution of time-series length in the HMHS and MIMIC-IV cohorts, respectively, while panels (c) and (d) show the distribution of time-series data density among candidemia-positive cases in HMHS and MIMIC-IV. Overall, the length and density of available time-series data differed between the HMHS and MIMIC-IV cohorts.

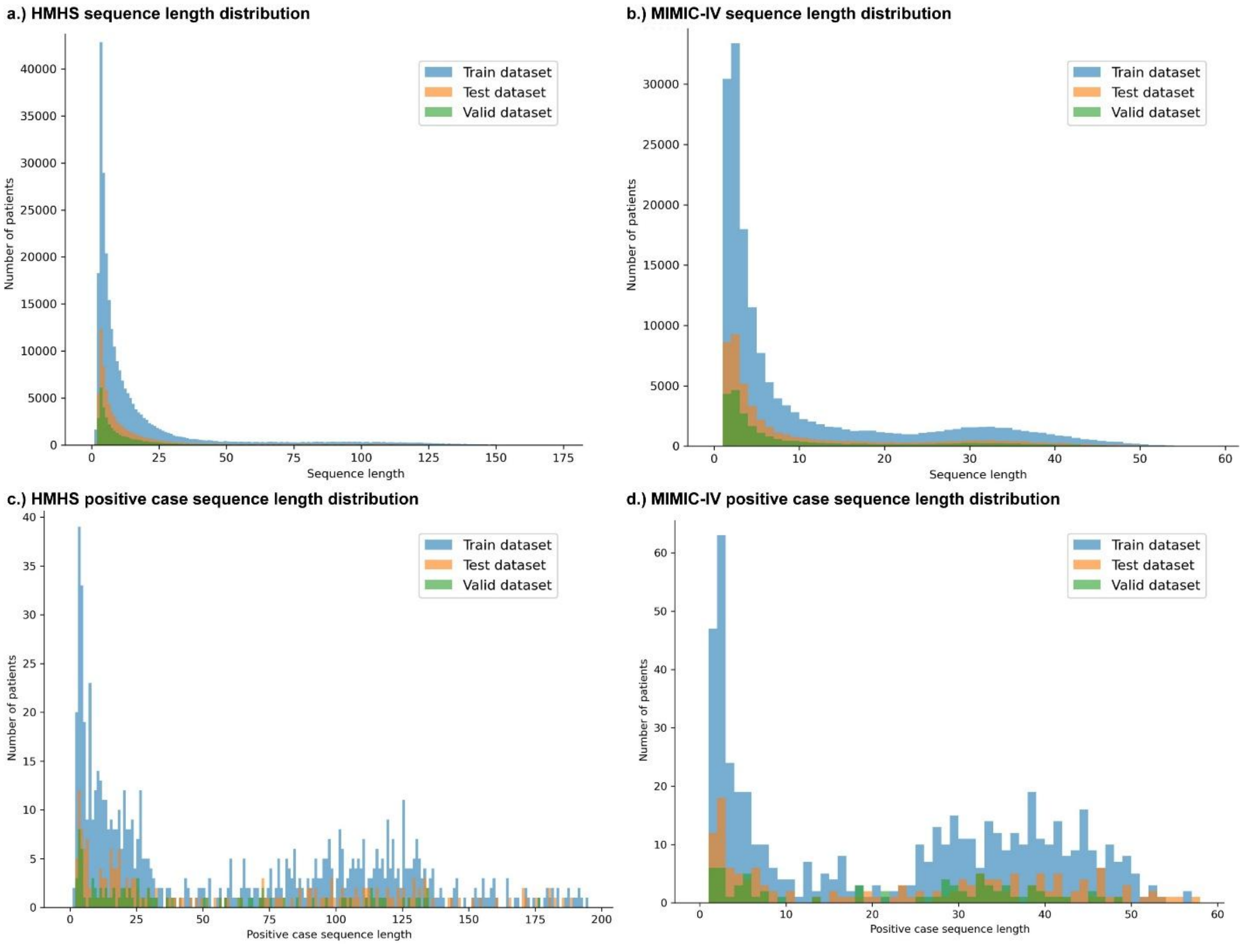

**Supplementary Figure S3. Receiver operating characteristic and precision–recall curves for candidemia prediction models in the HMHS and MIMIC-IV cohorts**

Panels (a) and (b) show ROC curves for the HMHS and MIMIC-IV datasets, respectively, comparing logistic regression (LR), LightGBM (LGBM), PyTorch_EHR, and transfer learning–based PyTorch_EHR (MIMIC-IV only). Panels (c) and (d) show corresponding precision–recall curves, highlighting model performance in the setting of extreme class imbalance. The horizontal dashed lines indicate the baseline precision corresponding to the observed candidemia prevalence in each dataset. Across both cohorts, PyTorch_EHR demonstrated superior discrimination compared with conventional machine learning models, particularly in terms of area under the precision–recall curve (AUPRC), reflecting improved identification of true candidemia cases in a rare-event setting.

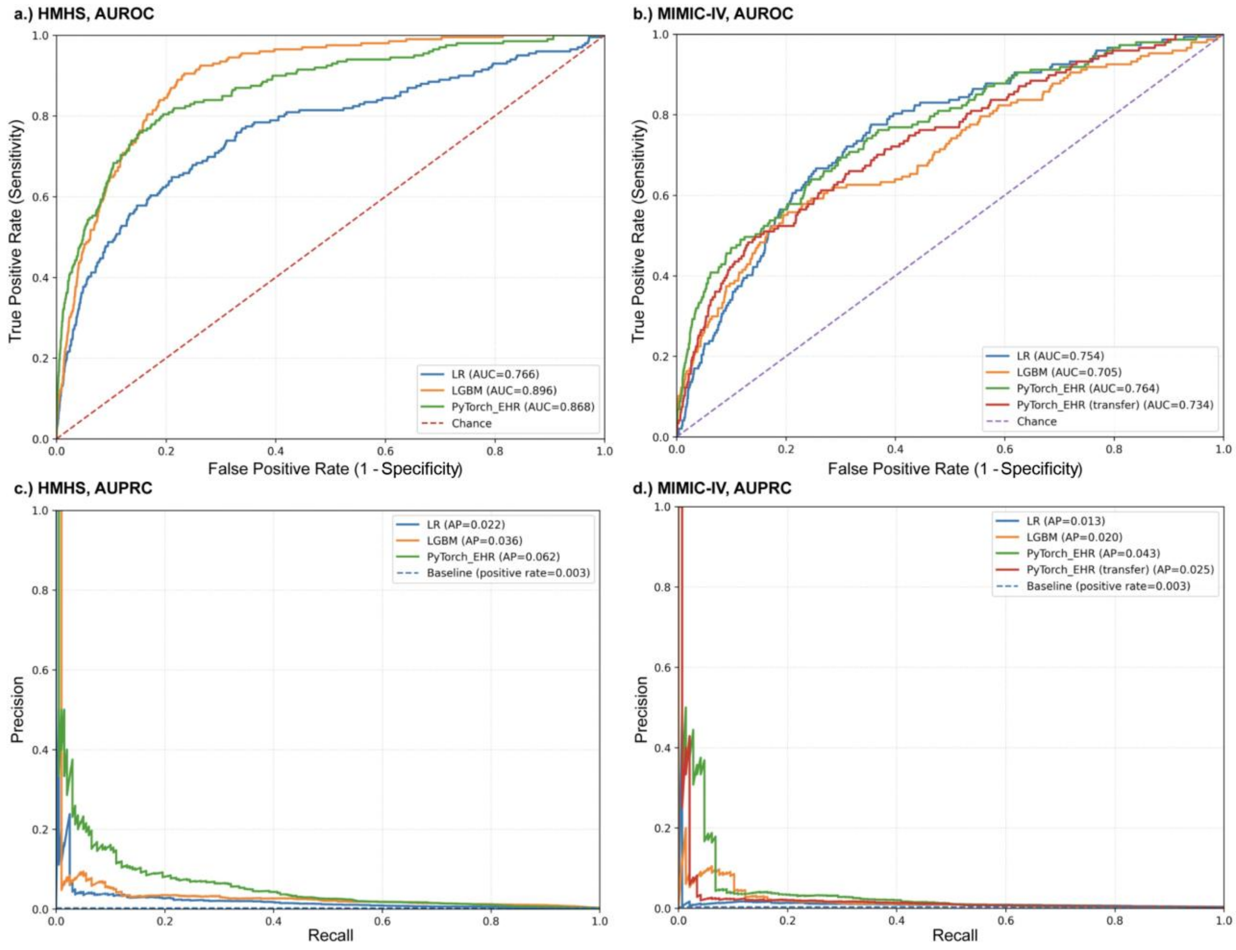

**Supplementary Table S4: Model performance for 30 days mortality prediction model (HMHS and MIMIC-IV)**

| | HMHS | | MIMIC-IV | |
|---|---|---|---|---|
| | AUROC | AUPRC | AUROC | AUPRC |
| PyTorch_EHR | 0.891 | 0.422 | 0.821 | 0.379 |
| PyTorch_EHR (Transfer learning from HMH data) | - | - | 0.826 | |

HMHS, Houston Methodist Hospital System; AUROC, area under the receiver operating characteristic curve; AUPRC, area under the precision–recall curve

**Supplementary Table S5: Diagnostic performance of the candidemia prediction model in the HMHS and MIMIC-IV cohorts**

| Cohort | Operating point | Sensitivity | Specificity | PPV | NPV |
|---|---|---|---|---|---|
| **HMHS** | Specificity 95% | 0.508 | 0.950 | 0.026 | 0.999 |
| | Sensitivity 90% | 0.905 | 0.581 | 0.006 | 1.000 |
| **MIMIC-IV** | Specificity 95% | 0.272 | 0.950 | 0.017 | 0.998 |
| | Sensitivity 90% | 0.905 | 0.307 | 0.004 | 0.999 |

HMHS, Houston Methodist Hospital System; PPV, positive predictive value; NPV, negative predictive value.

Operating points were defined based on prespecified targets for specificity (95%) or sensitivity (90%). Sensitivity, specificity, PPV, and NPV were calculated using 7-day candidemia as the outcome.

Given the low prevalence of candidemia (~0.3%), PPV and NPV values reflect the expected impact of class imbalance in rare-event prediction.

**Supplemental Table S6: Distribution of candidemia cases across treatment recommendation categories for one-step and two-step models**

| HMMS | | | | |
|---|---|---|---|---|
| | Treatment recommended | Treatment or close monitoring is considered | Recommendation can not be made by model | No Treatment recommended |
| One-step model | 101/3,898 (2.6%) | - | 78/28,171 (0.3%) | 20/44,295 (0.0%) |
| Two-step model | 101/3,898 (2.6%) | 20/2,818 (0.7%) | 58/25,353 (0.2%) | 20/44,295 (0.0%) |
| **MIMIC-IV** | | | | |
| | Treatment recommended | Treatment or close monitoring is considered | Recommendation can not be made by model | No Treatment recommended |
| One step model | 40/2,344 (1.7%) | - | 92/28,171 (0.3%) | 15/14,151 (0.1%) |
| Two step model | 40/2,344 (1.7%) | 28/2,974 (0.9%) | 64/26,765 (0.2%) | 15/14,151 (0.1%) |

HMHS, Houston Methodist Hospital System

The total number of candidemia cases was 199 in the HMHS cohort and 147 in the MIMIC-IV cohort.

**Supplementary Figure S4: Feature importance of the LightGBM models for candidemia prediction in the HMHS and MIMIC-IV cohorts.**

SHAP (SHapley Additive exPlanations) summary plots are shown for the LightGBM models trained on the (a) HMHS dataset and (b) MIMIC-IV dataset. Each point represents an individual event-level prediction, with the x-axis indicating the SHAP value (i.e., the contribution of each feature to the model output). Positive SHAP values indicate an increased predicted risk of candidemia, whereas negative values indicate a decreased risk. Features are ranked by overall importance from top to bottom. Color represents the feature value, with red indicating higher values and blue indicating lower values. Features were derived from structured electronic health record data, including laboratory measurements, diagnosis codes, procedure codes , and medication orders. TPN, total parenteral nutrition; EEG, electroencephalogram

**a.) HMHS**

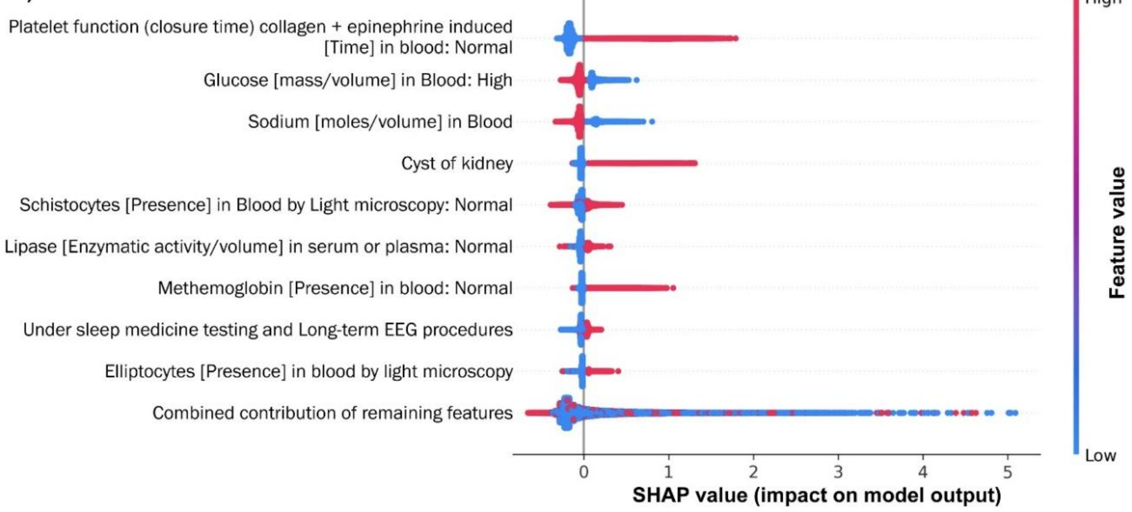


**b.) MIMIC-IV**

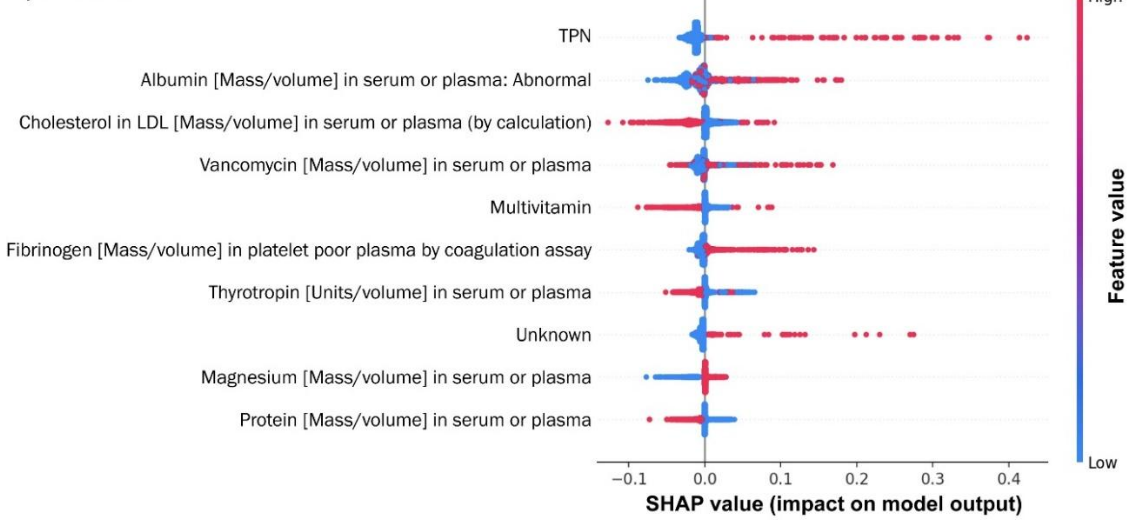

**Supplemental Table S7: Feature importance derived from integrated gradients for PyTorch-EHR candidemia prediction models in the HMHS and MIMIC-IV cohorts**

| HMHS | | MIMIC-IV | |
|---|---|---|---|
| **Feature** | **Mean \|IG\|** | **Feature** | **Mean \|IG\|** |
| Bacteria identified in blood by culture (Positive) | 0.0171 | Fat emulsion, intravenous | 0.0060 |
| Bacteria identified in blood by culture (Negative) | 0.0102 | Unspecified mycosis | 0.0044 |
| Adult hypertrophic pyloric stenosis | 0.0059 | Not Hispanic or Latino (Ethnicity) | 0.0038 |
| Hispanic or Latino (Ethnicity) | 0.0055 | Other | 0.0037 |
| Unspecified mycosis | 0.0053 | Hispanic or Latino (Ethnicity) | 0.0032 |
| Male | 0.0043 | Declined | 0.0030 |
| Severe sepsis with septic shock | 0.0039 | AGE_90 | 0.0026 |
| Declined | 0.0039 | Unavailable | 0.0023 |
| White (race) | 0.0036 | Unspecified hydronephrosis | 0.0021 |
| Unavailable | 0.0033 | AGE_80 | 0.0017 |
| Acute kidney failure, unspecified | 0.0031 | Male | 0.0015 |
| Arabic (Race) | 0.0027 | Triglyceride in serum or plasma | 0.0015 |
| AGE_100 | 0.0024 | AGE_30 | 0.0014 |
| Spanish (Language) | 0.0024 | Introduction of nutritional substance into peripheral vein | 0.0014 |
| Female | 0.0024 | Native American (Race) | 0.0014 |
| Asian (Race) | 0.0023 | Hydrocortisone | 0.0013 |
| Cellulitis of left lower limb | 0.0022 | Piperacillin–tazobactam | 0.0012 |
| AGE_20 | 0.0021 | AGE_40 | 0.0012 |
| Fever, unspecified | 0.0021 | Introduction of nutritional substance into central vein | 0.0012 |
| Altered mental status, unspecified | 0.0020 | Lipase in serum or plasma | 0.0011 |

HMHS, Houston Methodist Hospital System; IG, Integrated Gradients

Variables labeled as “Declined,” “Unavailable,” or “Other” indicate missing, refused, or non-specified information in the electronic health record and should be interpreted as proxies for clinical context rather than direct biological risk factors. Intravenous fat emulsions and procedural codes related to nutritional administration reflect exposure to parenteral nutrition and vascular access.

**Legend:** This table shows the top 20 features ranked by mean absolute integrated gradients (Mean |IG|) from PyTorch-EHR models developed to predict candidemia in the Houston Methodist Hospital System (HMHS) and MIMIC-IV cohorts. Mean |IG| reflects the average magnitude of each feature's contribution to the model output across patients, with larger values indicating greater overall importance. Features are listed separately for each cohort to highlight similarities and differences in influential clinical, demographic, laboratory, medication, and procedural variables.
